# Outcome Prediction Models for Critically Ill Patients Using Small Routine Laboratory Datasets

**DOI:** 10.64898/2026.04.26.26351758

**Authors:** Xiang Cao, Jianguo Hou, Xinhang Wei, Qi Wang

## Abstract

We present a suite of foundational, outcome prediction models for critically ill patients, developed using readily available, routine blood tests and advanced machine learning techniques. The input data of the models includes complete blood counts (CBCs), metabolic panels, and additional biomarkers that assess liver and kidney function, coagulation status, and cardiac injury. The output yields the predicted outcome at a given future horizon. For diagnoses, the length of the future horizon is set to zero while it is set to a fixed time interval for prognoses. The training dataset in this study comprises clinical data from 332 ICU patients, augmented with 200 synthetic samples generated via a conditional diffusion model. Generative machine learning–based data imputation and augmentation approaches yielded modest gains in predictive accuracy. However, substantial performance improvements were achieved through additional methods, including dimensionality and order reduction, SHAP-based feature importance analysis, and a novel time-series-to-image encoding strategy that enables the use of image-based classifiers for temporal clinical data. Principal component analysis–based order reduction produced measurable gains in outcome prediction, while the time-series-to-image encoding proved particularly effective in mitigating small-data limitations common in clinical research. Across all evaluation metrics—accuracy, precision, recall, F1 score, and AUROC—the prognostic models achieved performance exceeding 85%, with some models attaining AUROC scores above 90%. We innovated a new model-ensemble approach to optimize the predictive outcome. This ensemble modeling approach improves the overal prediction, pushing all assessment metrics over 90%. This work establishes a robust and interpretable AI-enabled diagnostic and prognostic toolkit for outcome predictions in critically ill patients and demonstrates a scalable workflow for developing high-performing models from sparse healthcare datasets. The proposed framework is readily deployable in ICU environments with routine blood testing capabilities and serves as a foundation for future integration into digital twin systems for critical care.

## 1 Introduction

Routine laboratory indicators, consisting of routine blood counts, liver and kidney function, coagulation function, and cardiac injury indicators, can be readily tested during a visit to the doctor’s office (see Table 1.1) [10, 12]. They provide key biochemical indicators that reflect the internal physiological and metabolic state of the patient and thus play crucial roles in the clinical decision-making process. They offer insights into various physiological and metabolic functioning components in the blood, such as cells, ions, sugars, lipids, hormones, proteins, and enzymes. As the body’s physiological and pathological states fluctuate over time, these measurements change accordingly, making them indispensable indicators for monitoring patient’s health state and inferring related potential abnormalities.

**Table 1.1:** Routine Biochemical Indicators.

|  |  |
| --- | --- |
| Routine blood counts | WBC, NEUT, NEUT%, LYM, LYM%, MONO, BASO, EOS, PCT, RBC, Hb, PLT |
| Liver and kidney function | BUN, ALT, AST, ALP, GGT, TBIL, DBIL, IBIL, TSBA, ALB, PA, CHE,<br>SCR, GFR, LDH, UA, Na, K, Ca, P, Mg, Cl |
| Coagulation function | PT, APTT, TT, FIB, FDP, D-Dimer |
| Cardiac injury markers | MYO, hs-TnT, CK-MB, BNP, CRP |

Upon patient admission, physicians rely on routine biochemical indicators to assess the patient’s homeostasis, the functionality of vital organs, relevant abnormalities and related disease severity. In modern medicine, these indicators are fundamental to medical diagnosis, intervention, and prognosis, guiding clinical decision-making and treatment design. Experienced physicians can extract or interpret collective features based on these indicators to predict patient outcomes in the ICU and implement timely medical interventions based on their many year’s experience and domain knowledge. In general, the more experience a physician has, the more accurate their predictions tend to be.

However, the scarcity of highly experienced physicians in ICU settings presents a major challenge to many communities. Junior physicians, due to limited clinical experience and the complex indications of the indicators, often struggle with making timely, appropriate, or accurate medical decisions. The intricate nature of ICU patient management necessitates a quick, intelligent decision-support tool that can assist less experienced physicians to accurately predict biochemical and physiological trends of the underlying patient’s based on available patient records and make appropriate medical diagnosis and prognosis.

In recent years, clinical predictive models based on routine biochemical indicators have gained significant attention in medical research and clinical implementations [6]. These models aim to forecast the onset of diseases, progression, and prognosis by analyzing patient biochemical profiles up to date and map them to patient’s health states [8]. Current research primarily focuses on predicting mortality rates across different populations, identifying risk factors associated with higher or lower survival rates, and distinguishing patient groups with better or worse prognoses [15, 16].

Despite the potential, many existing models suffer from poor quality and a lack of external validation, limiting their clinical applicability [7,8] Therefore, there is an urgent need for a more accurate, dynamic, and personalized predictive model that utilizes available spatial-temporal data of ICU patients’ simple biochemical indicators. Such a model could not only enhance the assessment of disease, diagnosis, and prognosis but also predict the trend of the biochemical indicators with more accurate trajectories, thereby improving clinical decision-making and patient care management. This advancement would mark a significant step toward precision medicine, ultimately improving patient outcomes and enhancing the quality of healthcare in general.

In this study, we aim to develop clinically implementable foundational diagnostic and prognostic models for critically ill ICU patients, leveraging readily available, routine biochemical indicators. The objectives are twofold: (1) to provide accurate diagnosis and prognostic assessments for ICU patients robustly, and (2) to demonstrate the feasibility of constructing foundation models using a dataset derived from a relatively small patient cohort. The specific biochemical indicators used in this study are listed in Table 1.1. The model is designed to generate patient-specific outcomes (diagnoses and prognoses) based on each individual’s simple laboratory record accumulated during hospitalization. Ultimately, it is intended to serve as an intelligent clinical decision support tool, aiding physicians in managing critically ill patients.

Data driven modeling using machine learning encompasses two indispensable ingredients: 1) good quality data and 2) effective machine learning models suitable for the intended task. In studies involving clinical data, a perennial issue is the size of the dataset. Usually they are small compared to other machine learning tasks in AI. In this study, we aim to address these two issues directly. We begin by implementing a suite of machine learning models to preprocess the data, with a particular focus on imputing missing values in a clinically reasonable and statistically sound manner. Once a complete dataset is obtained, we develop a range of classification models to predict patient outcomes in a fixed-length horizon, categorizing individuals into survival or deceased groups based on their historical records. To identify the most influential biochemical indicators, we employ reduced-order modeling and SHAP (SHapley Additive exPlanations) analysis, allowing for interpretable variable selection. Outcome prediction models are then built using these reduced sets of features. In parallel, we apply time-series-to-image encoding techniques to transform longitudinal patient data into image representations, enabling classification via image recognition models. All models are rigorously benchmarked and compared to establish a robust suite of outcome classifiers.

To enhance prediction performance in small-data settings, we explore data augmentation techniques using generative models to create synthetic samples. At the same time, we adopt dimensionality reduction methodologies to improve the ratio of observations to features, thereby strengthening classification performance. A variety of methods are examined in both directions, and each yields improved results. When several models demonstrate strong performance while excelling in different evaluation metrics, including accuracy, precision, F1 score, recall, and AUROC, we develop a novel ensemble approach for outcome prediction. This ensemble modeling strategy integrates the strengths of the individual models to produce a predictor that outperforms any single constituent model.

This AI-enabled modeling framework highlights a small subset of biochemical indicators, for which we provide a detailed discussion of their clinical significance and alignment with established medical knowledge. The framework, especially the modeling strategy applies to any EHR dataset of any size, making it truly a foundation model for outcome prediction derived from a small dataset. Collectively, the ensemble of models constitutes a comprehensive toolkit for ICU prognostication and offers a modular foundation for integration into future digital twin technologies for targeted diseases such as cancer, diabetes, or cardiovascular conditions.

## 2 Data Acquisition

This was a retrospective cohort study. The study site was in the Department of Emergency and Intensive Care Unit at Sichuan Provincial People’s Hospital, a comprehensive ICU equipped with 28 patient beds that admits approximately 1,200 patients annually. The research protocol was reviewed and approved by the Medical Ethics Committee of Sichuan Academy of Medical Sciences and Sichuan Provincial People’s Hospital (Approval No.: 2024-827). The ethics committee approved the waiver for informed consent. The study protocol is registered in the Chinese Clinical Trial Registry (https://www.chictr.org.cn/index.html) with the study registration number ChiCTR2500102396.

### 2.1 Inclusion and exclusion criteria

A retrospective collection of 332 ICU patients who met the inclusion criteria from January 2022 to December 2024 was obtained using the medical records information system of Sichuan Provincial People’s Hospital. Biochemical metabolic indicators from 4 to 7 days prior to the ICU outcomes were collected for these patients. The inclusion and exclusion criteria are given in Table 2.1.

**Table 2.1:** Inclusion and exclusion criteria.

| Inclusion criteria: | Exclusion criteria: |
| --- | --- |
| length of intensive care unit $\geq 4$ days; | APACHE II Scores $< 10$; |
| age $\geq 18$ years (adult patients); | Women who are pregnant or breastfeeding; |
| continuous biochemical data for at least 4 days; | Lack any outcome data; |
| APACHE II Scores at ICU admission $\geq 10$. | Participating in other clinical trials. |

### 2.2 Datasets

The data collected include the following:

- *Basic information of the patients*: age, gender, height, weight, APACHE II score, admission diagnosis, supplementary diagnosis, and discharge diagnosis.
- *Biochemical test data*: fasting blood test results collected at 6:00 AM daily during the patient’s hospitalization, including the indicators listed in Table 1.1.
- *ICU outcomes and final clinical outcomes of the patients*.

Mengjie Deng collected the data from 332 ICU patients, with records ranging from 4 to 33 days per patient and an average period of 8 days (Figure 2.1). For simplicity, each patient’s time series data were truncated to a maximum of 30 days, aligning all patients’ data based on their final recorded day. The dataset has totally 45 indicators, tabulated in Table 1.1.

**Figure 2.1:**
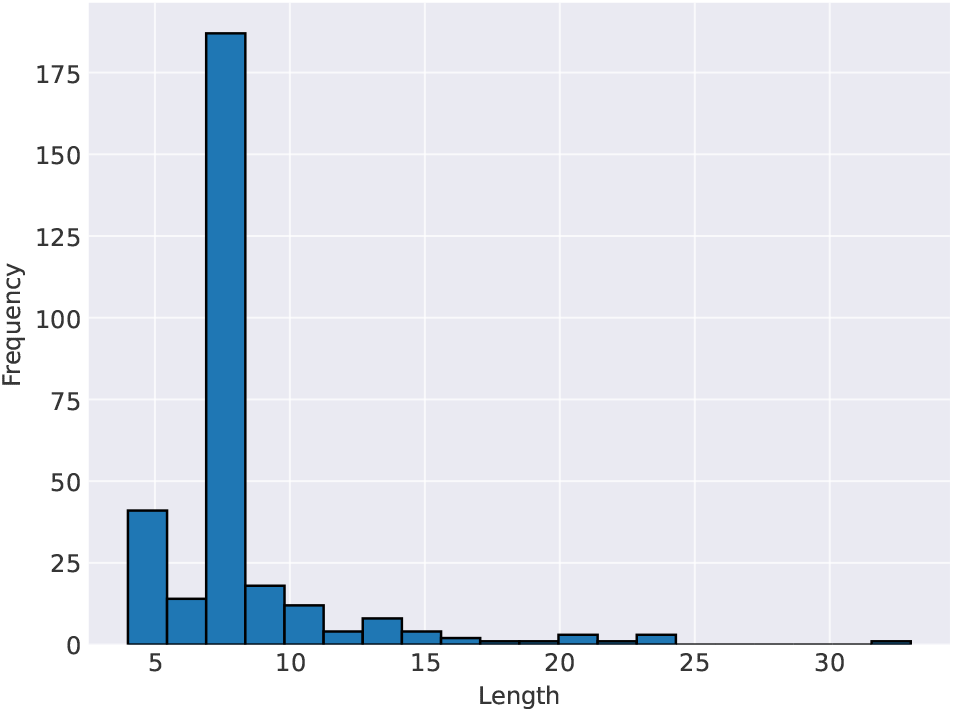
The number of recorded days for the participating ICU patients.

As shown in Table 2.3, most indicators have a missing data rate below 10%, although a few miss more than 90%. The average missing data rate is approximately 20%. To ensure the integrity of our data analysis, we removed five indicators with missing data rates greater than 60%, retaining 40 indicators in this study. The dataset was then split into the training, validation, and testing set in an approximate 8:1:1 ratio, resulting in 257 patients in the training set, 33 in the validation set, and 42 in the testing set. All details of the preprocessed datasets are presented in Table 2.2, while Table 2.3 provides comprehensive statistical information for each indicator.

**Table 2.2:**
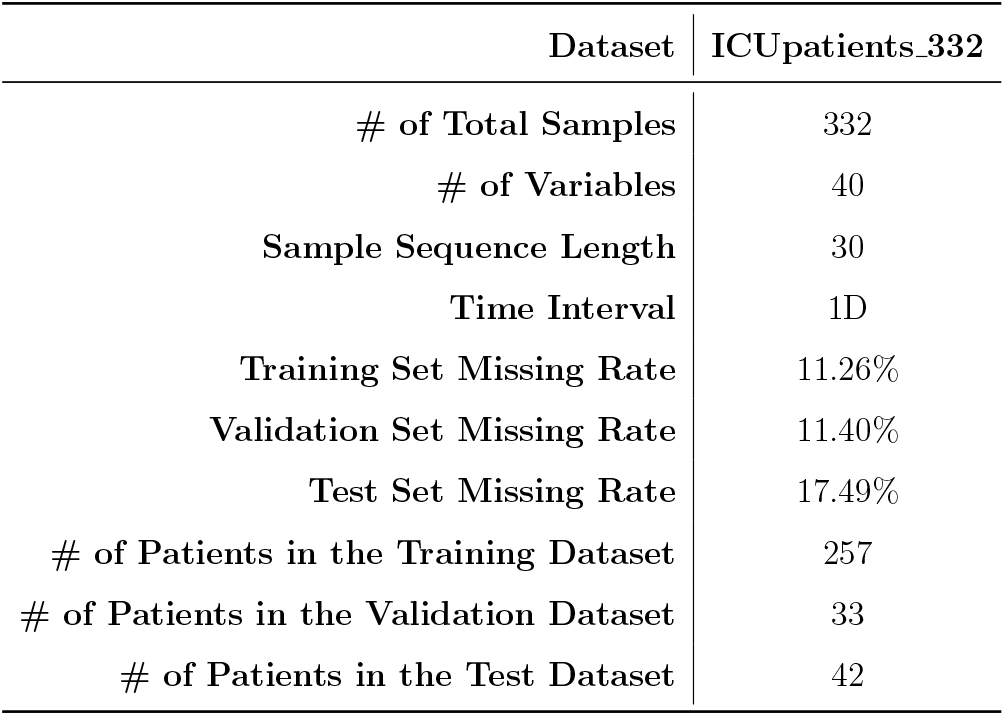
Details of the preprocessed datasets.

|  | Dataset | ICUpatients_332 |
| --- | --- | --- |
| # of Total Samples |  | 332 |
| # of Variables |  | 40 |
| Sample Sequence Length |  | 30 |
| Time Interval |  | 1D |
| Training Set Missing Rate |  | 11.26% |
| Validation Set Missing Rate |  | 11.40% |
| Test Set Missing Rate |  | 17.49% |
| # of Patients in the Training Dataset |  | 257 |
| # of Patients in the Validation Dataset |  | 33 |
| # of Patients in the Test Dataset |  | 42 |

**Table 2.3:**
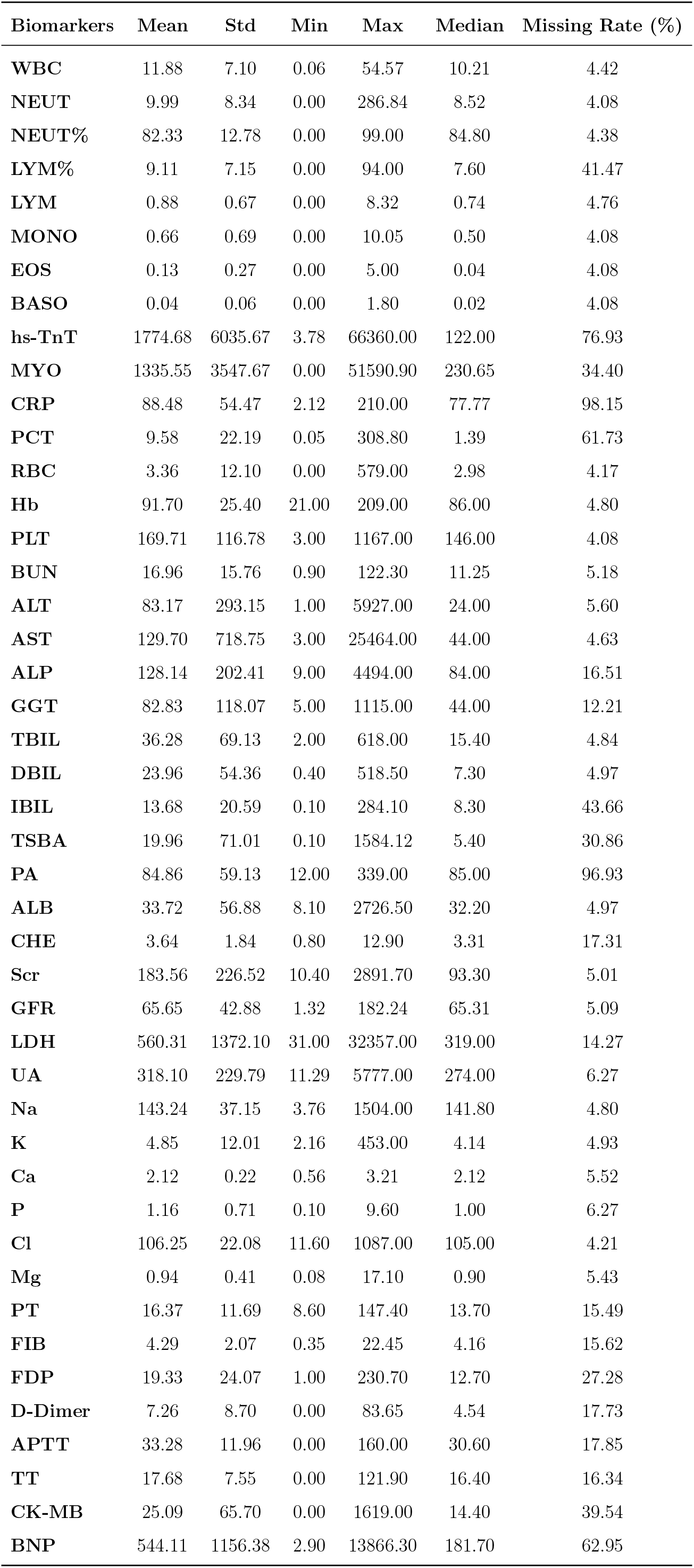
Statistical Information of Each Indicator.

## 3 Methods

Given that datasets collected or obtained from available databases normally contain certain percentage of missing data entries, it is imperative to make up the missing entries before the datasets can be used in any AI models. We therefore divide our study into two related tasks: data imputation and classification. Once a quality AI-ready dataset is available, there are usually multiple machine learning models to use to carry out the specified task. In this section, we explore a plethora of data imputation models for imputation tasks and a set of machine learning models for classification tasks to benchmark them on prognosis using routine biochemical indicators.

### 3.1 Imputation Task

A complete time series dataset can be denoted as 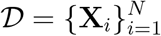. Each sample **X**_*i*_ ∈ ℝ^*L×K*^ is a matrix of observations, where *L* is the length of the time series, *K* is the number of variables, and *N* is the total number of time series in the dataset.

In practice, **X**_*i*_ is often incomplete. We can represent it as a combination of an observed part 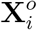 and a missing part 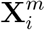. To formalize this, we use a binary observation matrix **M**_*i*_ ∈ {0, 1}^*L×K*^, where *m*_*l,k*_ = 1 if the value is observed and *m*_*l,k*_ = 0 if it is missing.

The goal of imputation is to train a model ℳ_*θ*_, parameterized by *θ*, to generate realistic estimates 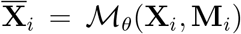 for the missing entries based on the observed data. The final imputed time series, 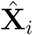, is then constructed by combining the original observed values with the model’s predictions:

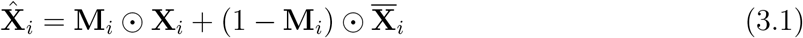

where ⊙ denotes element-wise multiplication. The first term preserves the original observed data, while the second term fills the missing positions with the values generated by the imputation model. The ultimate objective is that the reconstructed data 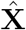 can closely approximate the true data distribution, thereby improving the performance of any downstream tasks.

#### 3.1.1 Data Imputation Models

We explore 9 imputation methods, including both machine learning and traditional methods.

The machine learning methods include the following:

- CSDI: Introduced by Tashiro et al. [21], this method uses conditional score-based diffusion models, which are trained to directly learn the conditional distribution of missing values based on observed data. The method also incorporates an attention mechanism to capture temporal and feature dependencies in the time series.
- GP-VAE: Developed by Fortuin et al. [11], the method contributes a novel VAE architecture for multivariate time series imputation, combined with a Gaussian Process (GP) prior to model temporal dynamics effectively.
- Transformer: Vaswani et al. [23] introduces the self-attention mechanism, which can process sequence data by paying attention to different positions within the sequence, leading to significant developments in natural language processing and time series fields.
- US-GAN: Miao et al. [19] integrates a classifier in the generative adversarial network to predict labels and provide feedback to the generator, guiding it to produce more accurate imputations. The model also introduces a temporal reminder matrix, which helps the discriminator better distinguish observed components from the imputed ones.
- Ensemble: Model ensemble is a machine learning technique where multiple models are combined to produce a stronger overall model. The expectation is that by combining the predictions of multiple models, the ensemble can achieve better performance than any single model alone.

The traditional methods consist of:

- Mean: This method fills in missing values by calculating the mean of the observed values in the time series.
- Median: This method fills in missing values by calculating the median of the observed values in the time series.
- Linear interpretation: This method fills in missing values by connecting the last observation before the missing data and the first observation after the missing data with a straight line.
- Locf(Last Observation Carried Forward): It fills in missing values by carrying forward the last observed value.

To benchmark the imputation methods, we use two commonly used metrics given below.

MSE (Mean Square Error):

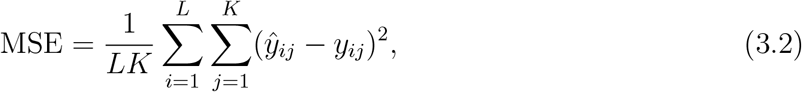

where *ŷ*_*ij*_ is the *i*-th predicted value of the *j*-th variable, *y*_*ij*_ is the corresponding value of the ground truth, *L* is the length of prediction and *K* is the number of indicators/variables.

MAE(Mean Absolute Error):

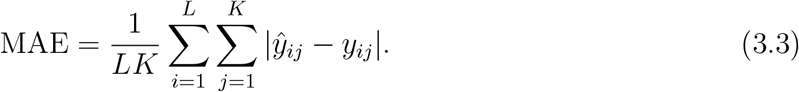

Table 3.1 summarizes performance of each imputation method where 10% of the observed values are randomly selected as ground truth in the testing set. For both the CSDI and GP-VAE models, the reported results represent the mean values over 100 samples. For the Ensemble model, we first apply the four pre-trained models to the training, validation, and testing sets, respectively, and then concatenate their outputs to form new datasets. Subsequently, we train a one-layer linear model on these new datasets. Essentially, this approach computes a weighted average of the four models, with the corresponding weights detailed in Table 3.2.

**Table 3.1:**
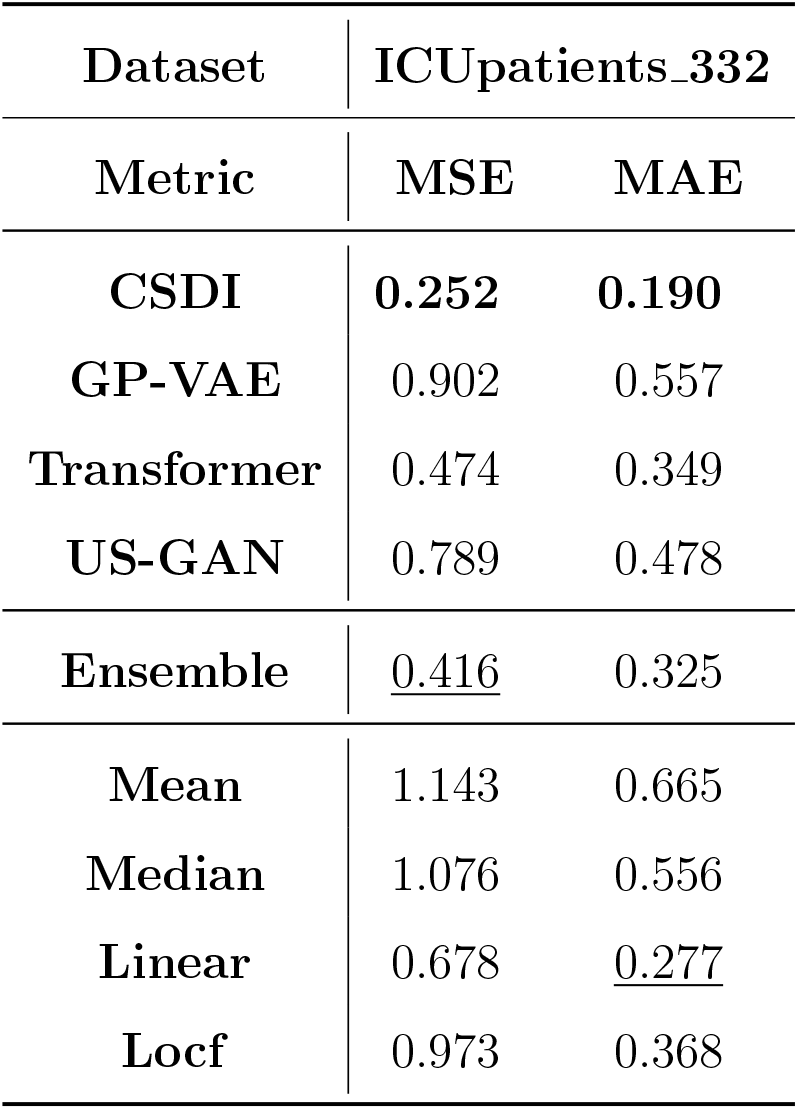
Performance of each imputation method with 10% missing values.

**Table 3.2:** The weights of each model in the Ensemble method.

| Dataset | ICUpatients_332 |
| --- | --- |
| **CSDI** | 0.20 |
| **GP-VAE** | 0.00 |
| **Transformer** | 0.36 |
| **US-GAN** | 0.43 |

The best-performing results in MSE and MAE in Table 3.1 are highlighted in bold, while the second-best ones are underlined. Among the assessed models, CSDI achieves the lowest MSE (0.252) and MAE (0.190), indicating the best overall performance. The ensemble method shows moderate improvements over some individual models (MSE=0.416, MAE=0.325), though it still falls short of CSDI’s results. The linear model gets the second place finish measured in MAE. Traditional imputation methods display higher errors, highlighting the advantages of machine learning-based methods in capturing complex temporal patterns while used in imputation. To illustrate the performance of the CSDI model, we visualize a single case focusing on the last 12 indicators in Figure 3.1, and present a comparison of four machine learning models in Figure 3.2.

**Figure 3.1:**
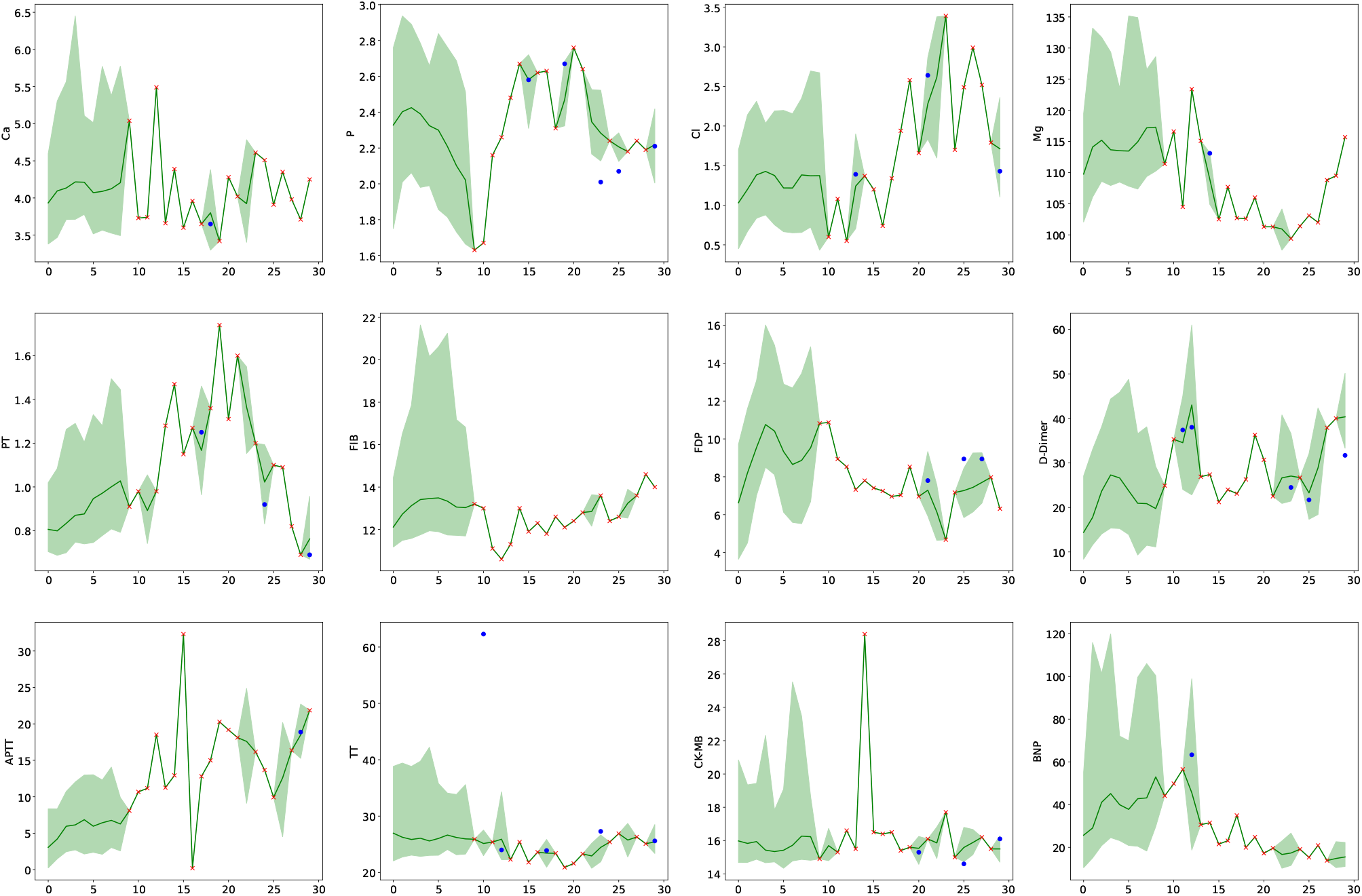
Visualization of probabilistic imputation results with 10% missing values for the last 12 features. The red crosses indicate observed values, while blue circles indicate ground-truth imputation targets. The green lines represent median imputations, and the shaded area represents the 90% confidence interval.

**Figure 3.2:**
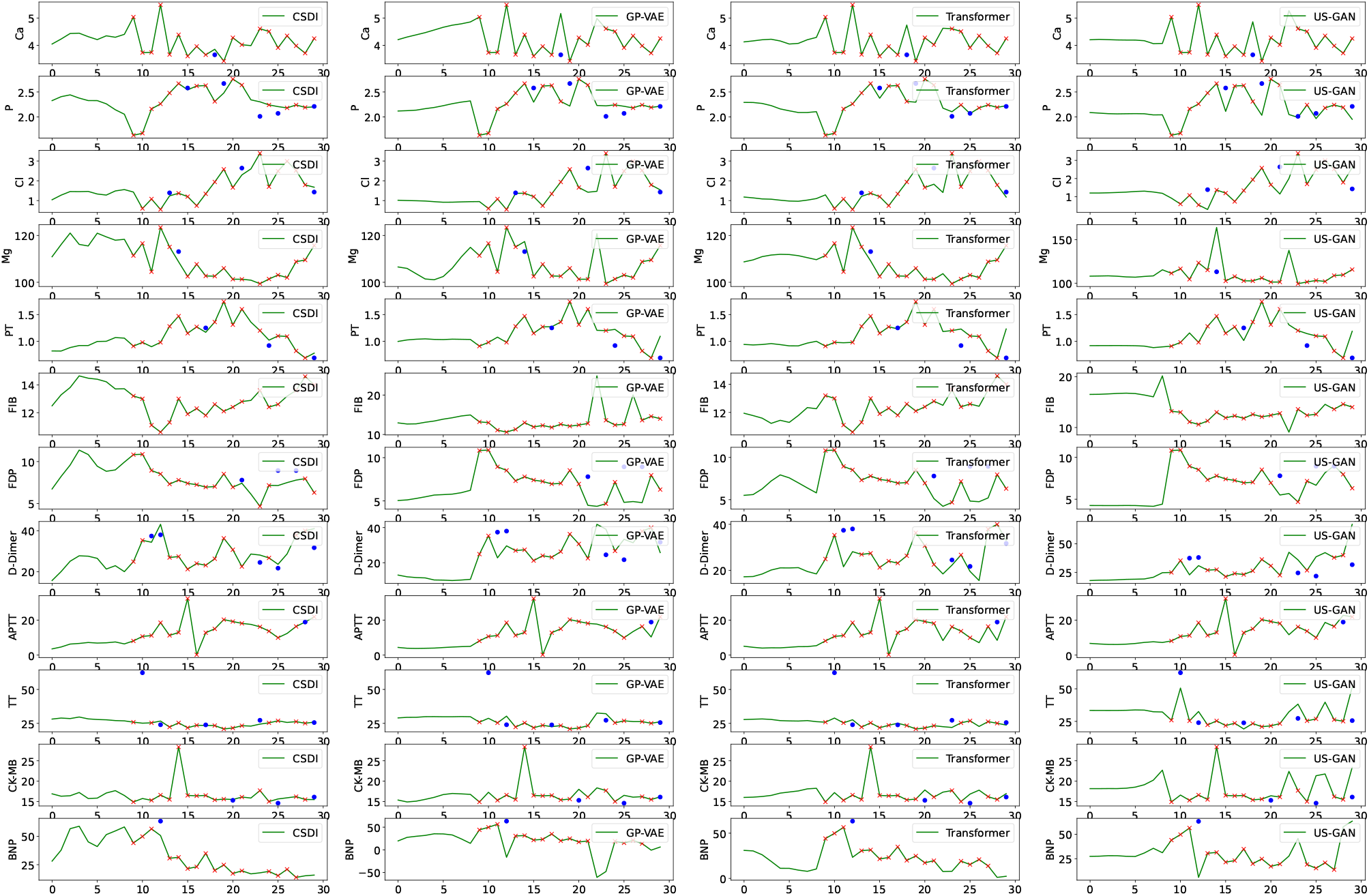
Comparison of four machine learning models (CSDI, GP-VAE, Transformer, and US-GAN) for time series imputation with 10% missing values. The red crosses indicate observed data points, blue circles indicate the ground-truth values, and the green line shows the predicted imputations.

### 3.2 Classification Task

Following the imputation step, the now complete time series dataset serves as the input for the prognostic classification task. Specifically, let 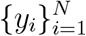 be the set of class labels, where each *y*_*i*_ represents the category of the *i*-th time series **X**_*i*_. In our work, label 1 indicates death, while label 0 indicates survival. Our goal is to train a classification model Φ, which takes the imputed time series 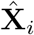 as an input and predicts the corresponding class label *ŷ*_*i*_.

Formally, for each reconstructed sample 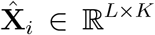, our classification model outputs a prediction:

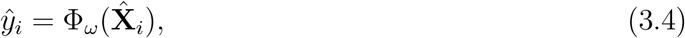

where *ω* denotes the learnable parameters of Φ. Our objective is to find parameters *ω* that minimize the classification loss (cross-entropy loss) over all samples:

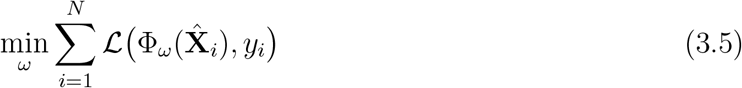

By leveraging the imputed time series, the classification model can better capture the underlying temporal patterns that were previously obscured by missing values. Hence, a more accurate imputation is expected to show better classification performance.

#### 3.2.1 Classification Models Based on TCN

After performing imputation in the clinically acquired dataset, we use the completed (i.e., AI-ready) dataset for the target binary classification. Specifically, we employ a CNN-based model, TCN.

- TCN: Bai et al. [2] use causal and dilated convolutions to capture long-range dependencies efficiently, often outperforming RNNs in accuracy and convergence speed.

In a binary classification setting, model performance is typically evaluated using a confusion matrix, given in Appendix. This matrix summarizes the counts of correct and incorrect predictions by comparing the ground truth labels with the model’s predictions. From this confusion matrix, several commonly used metrics are derived: Accuracy, Precision, Recall, F1-Score, and AUROC.

#### 3.2.2 Traditional Machine Learning Methods

We compared the classification performance of a series of traditional machine learning methods, including Random Forest [4], SVM [13], and XGBoost [5], with the previous TCN model in Table 3.3. All experiments were repeated 10 times using different random seeds, and the reported values represent the mean *±* standard deviation. As shown, the Random Forest model exhibits the best overall performance, achieving the highest scores in Accuracy, Precision, F1-score, and AUROC. These findings suggest that for classification tasks on small datasets, traditional methods can perform competently or even outperform deep learning approaches. At least for the dataset we have, the traditional models all outperform TCN!

**Table 3.3:** Comparison of classification performance between traditional methods and TCN.

| Methods | Accuracy | Precision | Recall | F1 | AUROC |
| --- | --- | --- | --- | --- | --- |
| **Random Forest** | **87.14±2.43** | **93.61±3.20** | 83.33±3.73 | **88.09±2.34** | **88.52±0.95** |
| **SVM** | 80.95±0.00 | 80.77±0.00 | 87.50±0.00 | 84.00±0.00 | 86.69±0.00 |
| **XGBoost** | 85.24±2.08 | 93.25±2.20 | 80.00±3.12 | 86.08±2.10 | 87.87±0.84 |
| **TCN** | 82.62±3.56 | 81.64±3.73 | **90.00±3.51** | 85.56±2.86 | 87.36±1.16 |

#### 3.2.3 Classification Models Based on Time-Series-to-Image Encoding

Given the rapid advances in imaging analysis using deep learning approaches, we next explore the time-series to image encoding approach to leverage the power of image analyzers in classification tasks. We introduce ViTST [17], which transforms irregularly sampled multi-variate time series into RGB images by visualizing variables as subplots within a fixed grid. We investigate three distinct encoding representations: line graphs (Figure 3.3), density plots (Figure 3.4), and bar charts (Figure 3.5). These generated images are then fed into a fine-tuned vision transformer or ResNet to capture temporal patterns. As presented in Table 3.4, all three encoding strategies outperform the TCN baseline, demonstrating the potential of leveraging advanced vision models for time series classification. Specifically, the Bar plot configuration yields the best overall performance, achieving the highest precision, and AUROC. Meanwhile, the Line plot excels in recall, underscoring the promise of this representation strategy.

**Figure 3.3:**
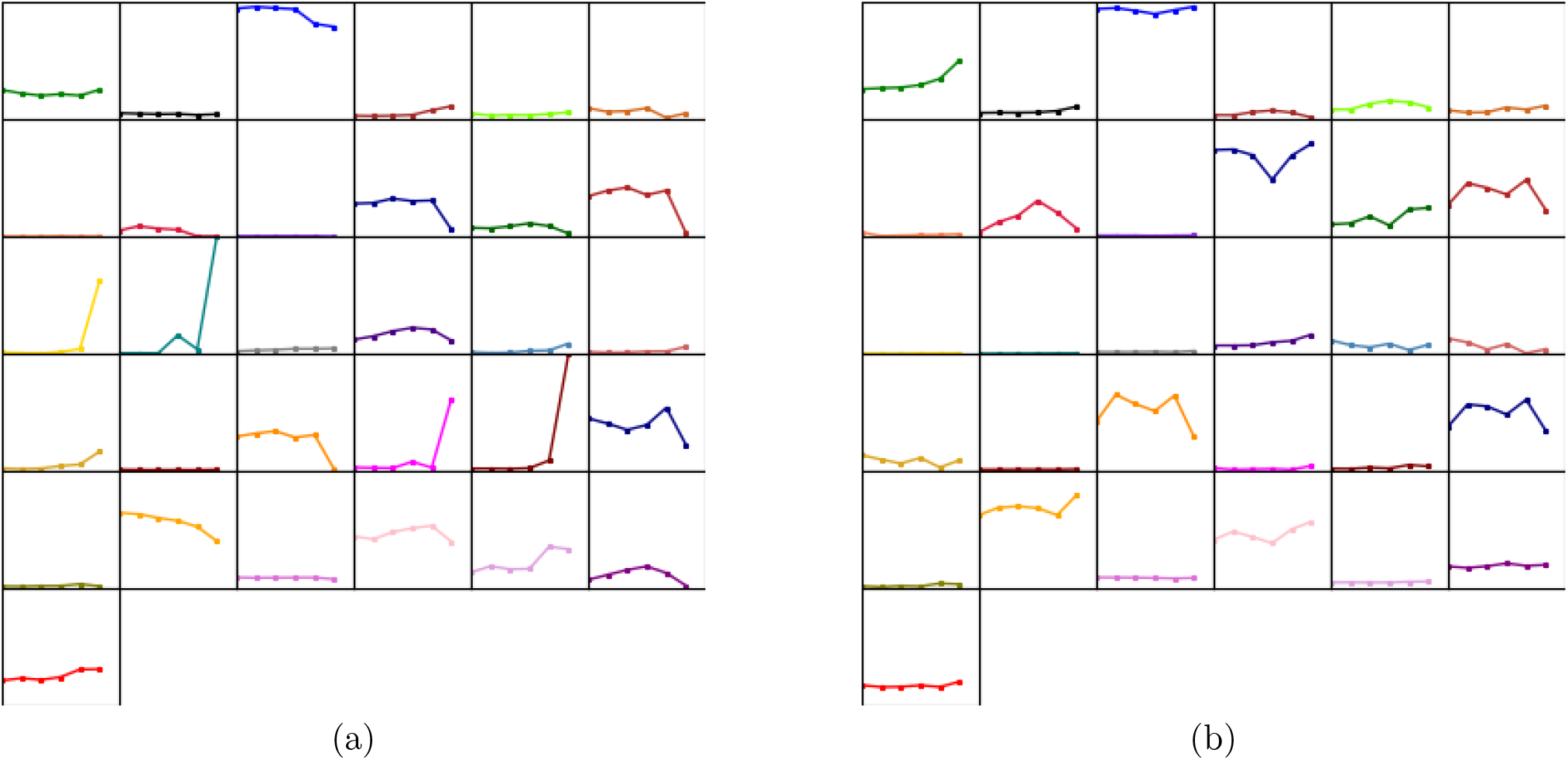
Two examples of how the original time series are converted into images and fed into the pre-trained classification model.

**Figure 3.4:**
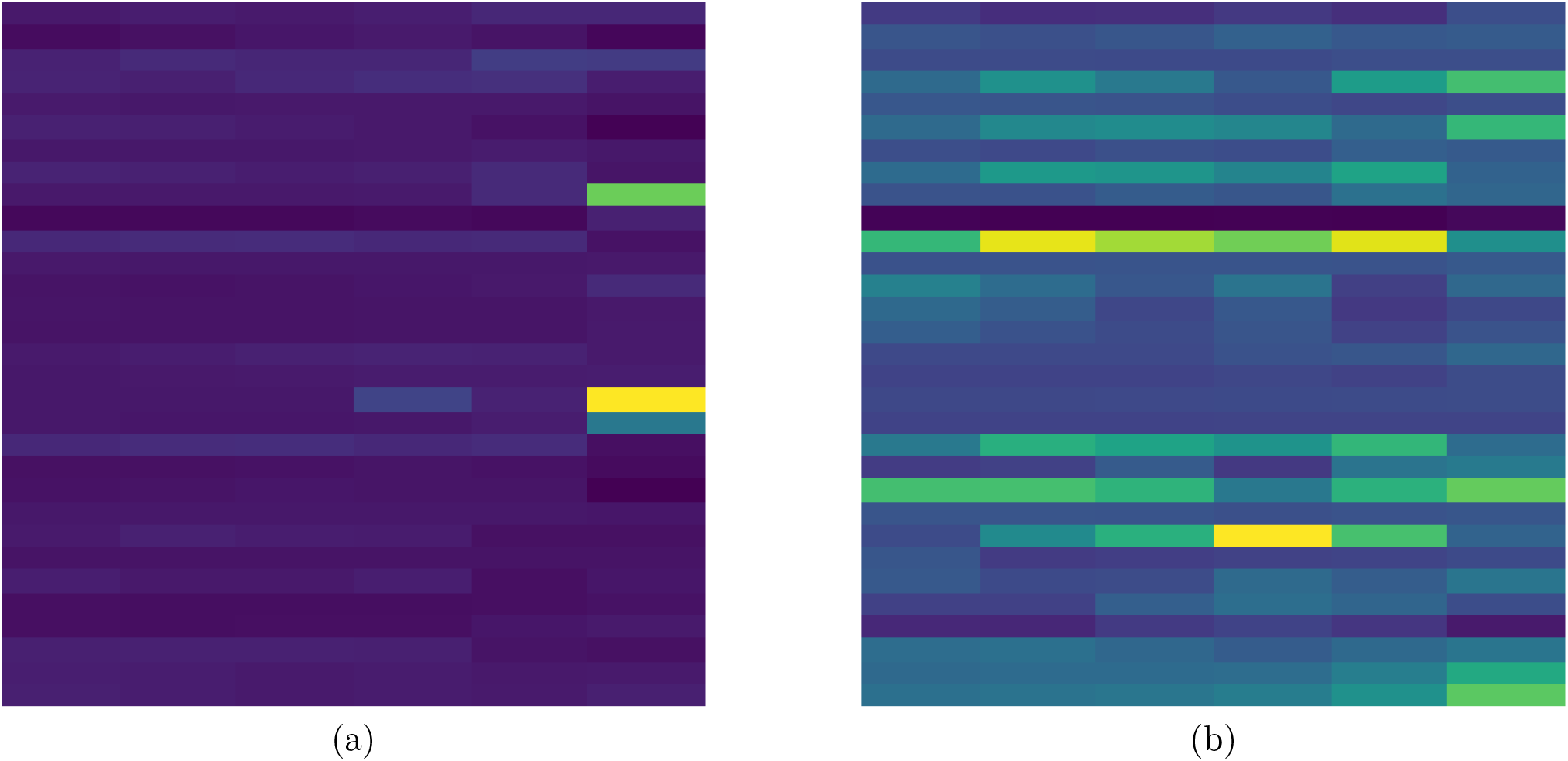
Density plots of two examples.

**Figure 3.5:**
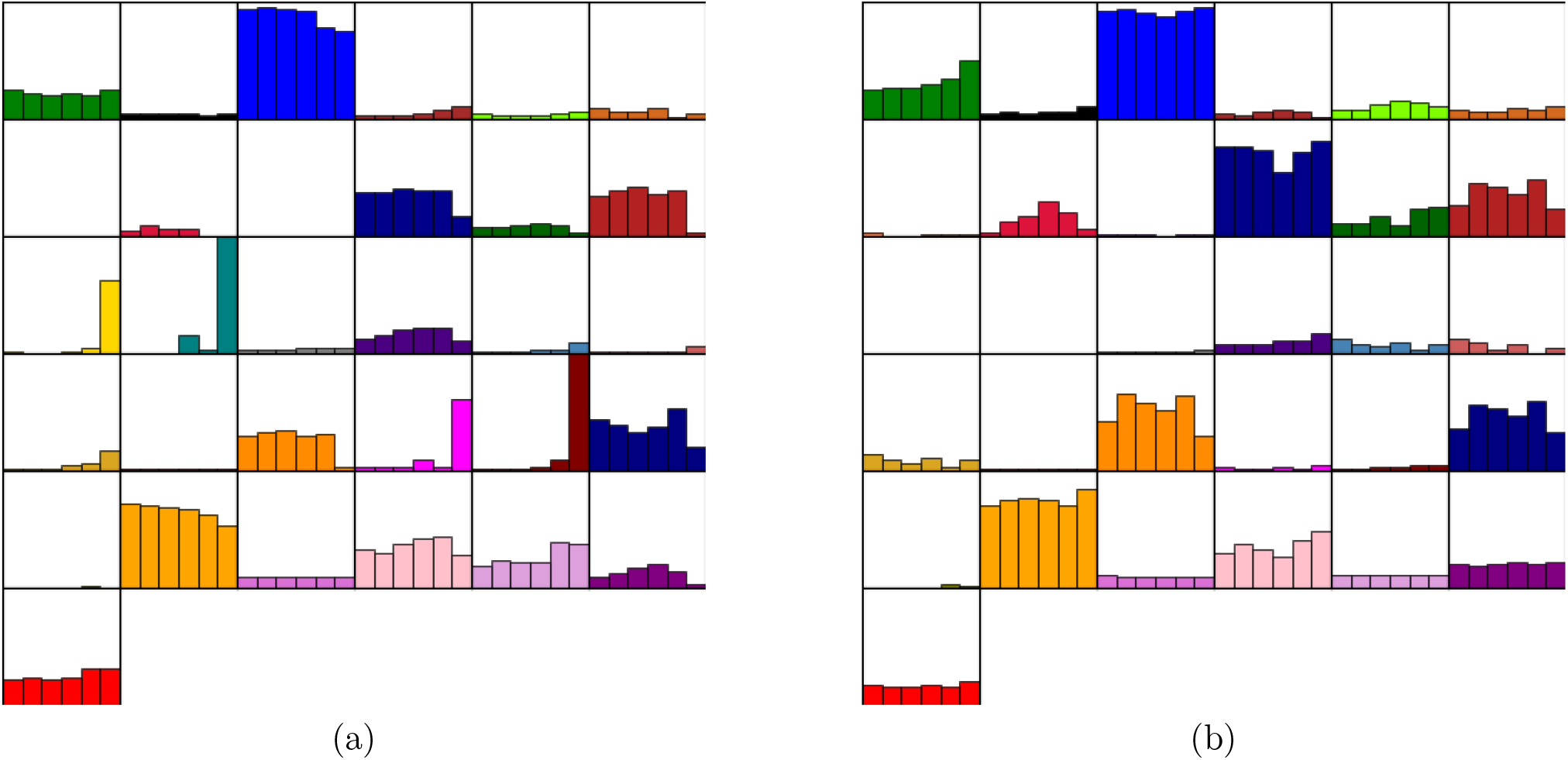
Bar-chart plots of two examples.

**Table 3.4:** Comparison of classification performance between ViTST and TCN.

| Methods | Accuracy | Precision | Recall | F1 | AUROC |
| --- | --- | --- | --- | --- | --- |
| **Line** | 81.43±4.97 | 85.17±4.87 | 82.08±7.92 | 83.34±4.81 | 87.08±3.04 |
| **Heatmap** | 80.00±3.72 | 81.05±2.58 | 85.00±7.50 | 82.78±3.96 | 86.30±2.07 |
| **Bar** | 80.00±5.35 | **89.61±4.98** | 74.58±13.88 | 80.29±7.59 | **88.31±1.57** |
| **TCN** | **82.62±3.56** | 81.64±3.73 | **90.00±3.51** | **85.56±2.86** | 87.36±1.16 |

Another advantage of the time-series-to-image encoding is that the time series does not need to be aligned perfectly from patient to patient. I.e., the time stamps in one patient’s time-series can differ from those in another patient’s. This is a particular appealing feature offered by this simple yet effective approach.

### 3.3 Data Augmentation through Synthetic Data Generation

We note that a dataset consisting of 332 patients is small for data-driven modeling via machine learning. However, it is well-known that a systematic clinical data collection is extremely difficult to warrant high rate of fidelity without missing entries in large quantities. Generative model used in data imputation such as the diffusion model gives us a plausible route to generate more synthetic data that obey the same distribution as those of the ground truth data. Consequently, we experiment with data augmentation techniques using a diffusion model to see if we can improve predictions of the prognostic models.

### 3.4 Conditional Diffusion Model

Classifier Guidance [9] leverages gradient information from a pre-trained classifier to guide the sampling process toward specific class conditions. We consider the stochastic differential equation (SDE) formulation of diffusion models, governed by the following forward process:

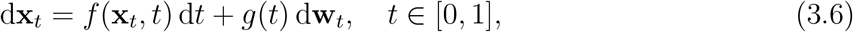

where *f* (**x**_*t*_, *t*) represents the drift term, *g*(*t*) the diffusion coefficient, and **w**_*t*_ a standard Brownian motion. Starting from a clean data sample **x**_0_ ∼ *p*_data_, this forward SDE gradually perturbs samples into a Gaussian distribution at *t* = 1.

The generative process is defined by the corresponding reverse-time SDE:

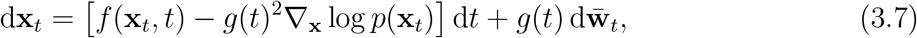

where *p*(**x**_*t*_) denotes the probability density of **x**_*t*_ and 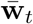 is the reverse-time Brownian motion. When generating samples conditioned on a label *y*, the reverse SDE is modified as follows:

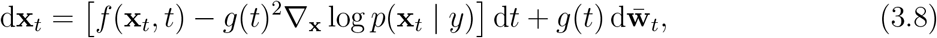

Applying Bayes’ rule to the conditional score function, we get

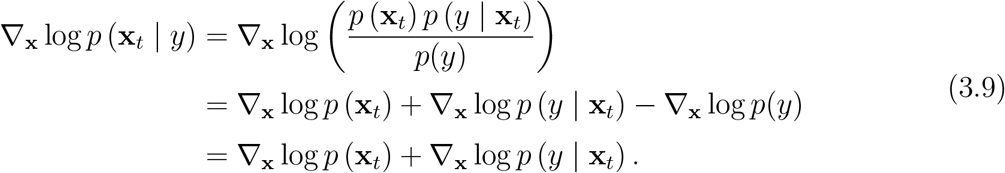

Thus, we need to train two parts. The first term, ∇_**x**_ log *p*(**x**_*t*_), is the unconditional score, which is learned during the diffusion model training. The second term, ∇_**x**_ log *p*(*y* | **x**_*t*_), is the gradient of a classifier and can be obtained from a neural network trained to approximate *p*(*y* | **x**_*t*_). Once both parts are available, we can use Eq. (3.8) to iteratively generate conditional samples.

We parameterize the unconditional score using a neural network *s*_*θ*_(**x**_*t*_, *t*). The objective is to match *s*_*θ*_(**x**_*t*_, *t*) with the true score ∇_**x**_ log *p*(**x**_*t*_) by minimizing:

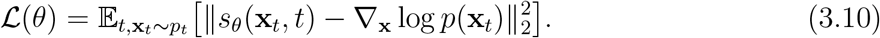

However, since the true score ∇_**x**_ log *p*(**x**_*t*_) is unknown, we utilize the property of Gaussian perturbations where **x**_*t*_ = **x**_0_ + *σ*(*t*)*ϵ*, with **x**_0_ ∼ *p*_data_ and *ϵ* ∼ 𝒩 (0, *I*). One can show (see [20]) that

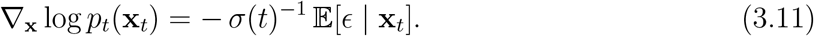

Substituting this into Eq. (3.10) yields the standard score-matching loss, which is equivalent to ℒ up to a constant. In practice, we minimize

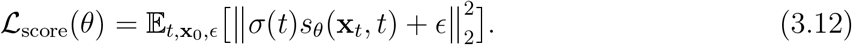

To control the influence of the condition during sampling, we introduce a guidance scale *λ >* 0 and modify Eq. (3.9) as

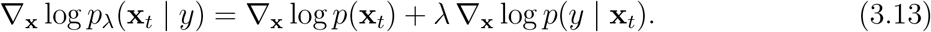

When *λ* = 0, the process ignores the label and reduces to unconditional sampling. As *λ* increases, the generation becomes increasingly biased toward samples that satisfy the condition *y*. However, one drawback of this approach is that it requires training an additional classifier *p*(*y* | **x**_*t*_).

Classifier-Free Guidance [14] is another method used in diffusion models that enables guided sample generation without an extra classifier. Instead, we use a single neural network 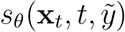 to handle both conditional and unconditional generation. The input 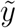 represents either the class label *y* or a null token ∅ indicating the absence of conditioning.

During training process, we randomly drop the condition with a fixed probability *p*_drop_ = 0.1. Specifically, for each training step, we sample

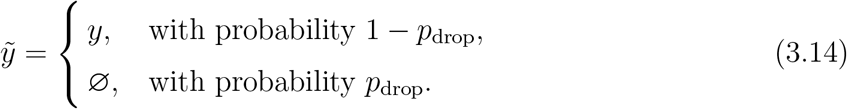

The same score-matching loss is then applied to 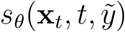:

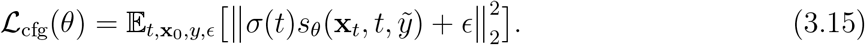

When 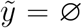, the network learns to approximate the unconditional score ∇_**x**_ log *p*(**x**_*t*_). When 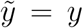, it learns the conditional score ∇_**x**_ log *p*(**x**_*t*_ | *y*). Thus, a single network learns both behaviors.

For the sampling process, we substitute Eq. (3.9) into Eq. (3.13) to remove the classifier term:

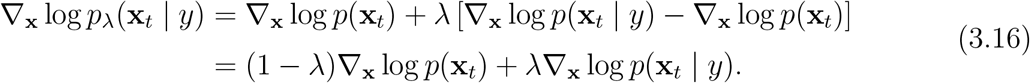

We then estimate these two scores by evaluating the same network with different conditional inputs:

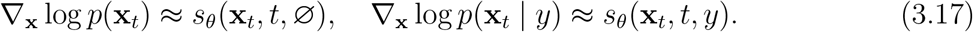

By plugging these neural network approximations into Eq. (3.16), we obtain the formulation used in our sampling process. Notably, when *λ >* 1, the sampler moves away from the unconditional distribution and strictly towards the class conditional distribution, thereby enhancing the relevance of the generated time series to the patient’s specific outcome.

In our experiments, we varied *λ* from 0 to 7.5, with the corresponding results summarized in Table 3.5. To better compare the imputation performance under different guidance scales, Figure 3.6 presents a representative case for PT and D-Dimer.

**Table 3.5:** Comparison of imputation results under different guidance scales *λ*.

| $\lambda$ | MSE | MAE |
| --- | --- | --- |
| 0 | 0.264 | 0.201 |
| 1 | 0.264 | 0.198 |
| 3 | **0.259** | **0.197** |
| 7.5 | 0.265 | 0.212 |

**Figure 3.6:**
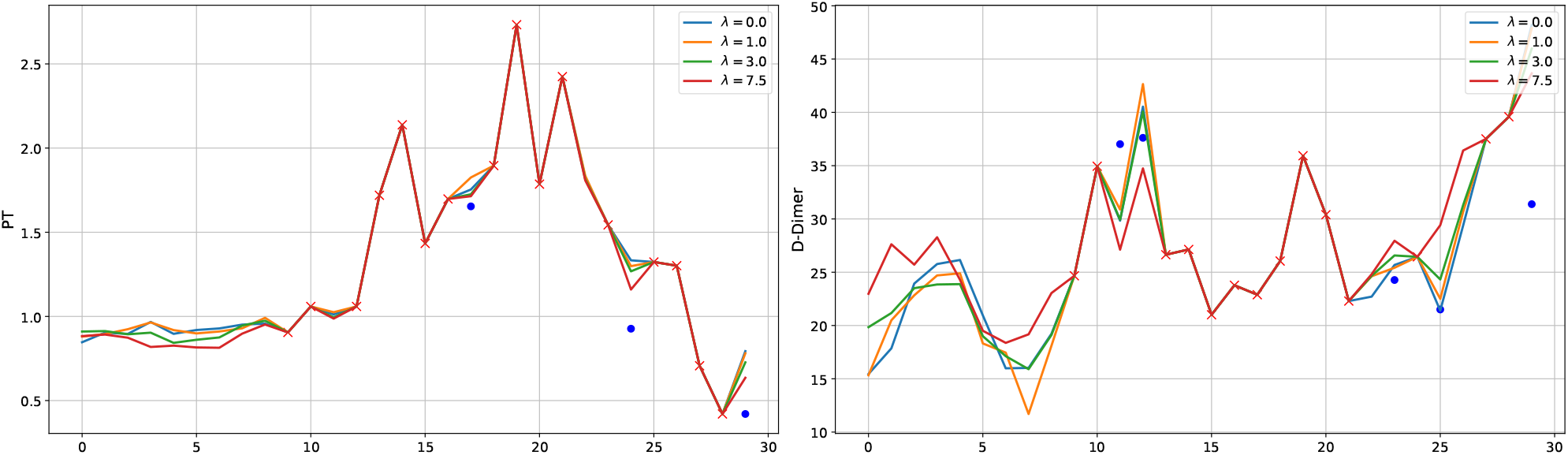
Comparison of PT and D-Dimer imputation results under different guidance scales *λ*, with ground-truth values shown as blue dots.

Based on the results in Table 3.5, we selected *λ* = 3 for the following analysis. It is important to note that when a patient data is entirely missing, the imputation process essentially becomes data augmentation. Given that the original dataset’s ratio of positive to negative samples is 9:11, we generated 90 additional positive and 110 additional negative samples accordingly. The resulting classification performance is summarized in Table 3.6. Notably, data augmentation leads to improvements in Accuracy, Precision, Recall and F1 score, while AUROC shows a slight decrease. Figure **??** provides a visual illustration of these results. The improvements range from a 0.47% increase in recall to a 3.17% increase in precision, highlighting the usefulness of data augmentation in enhancing overall classification performance.

**Table 3.6:** Comparison of classification performance on the original and augmented datasets.

| Dataset | Accuracy | Precision | Recall | F1 | AUROC |
| --- | --- | --- | --- | --- | --- |
| Original | 82.62±3.56 | 81.64±3.73 | 90.00±3.51 | 85.56±2.86 | 87.36±1.16 |
| Augmentated | 84.76±3.76 | 84.23±4.06 | 90.42±2.81 | 87.18±3.04 | 86.02±1.44 |
| Improvement | 2.59% | 3.17% | 0.47% | 1.89% | -1.53% |

We visualize two synthetic cases in Figure 3.7. Figure 3.8 illustrates a t-SNE visualization comparing the original and augmented data for each class. T-SNE [22] is a non-linear dimensionality reduction technique that maps high dimensional data into a low dimensional space (typically 2D or 3D) for visualization. It converts pairwise similarities into probability distributions and minimizes the KL divergence between the high and low dimensional spaces to preserve local structure. Notably, the two datasets merge closely, suggesting that the augmented data has effectively learned the original data’s distribution.

**Figure 3.7:**
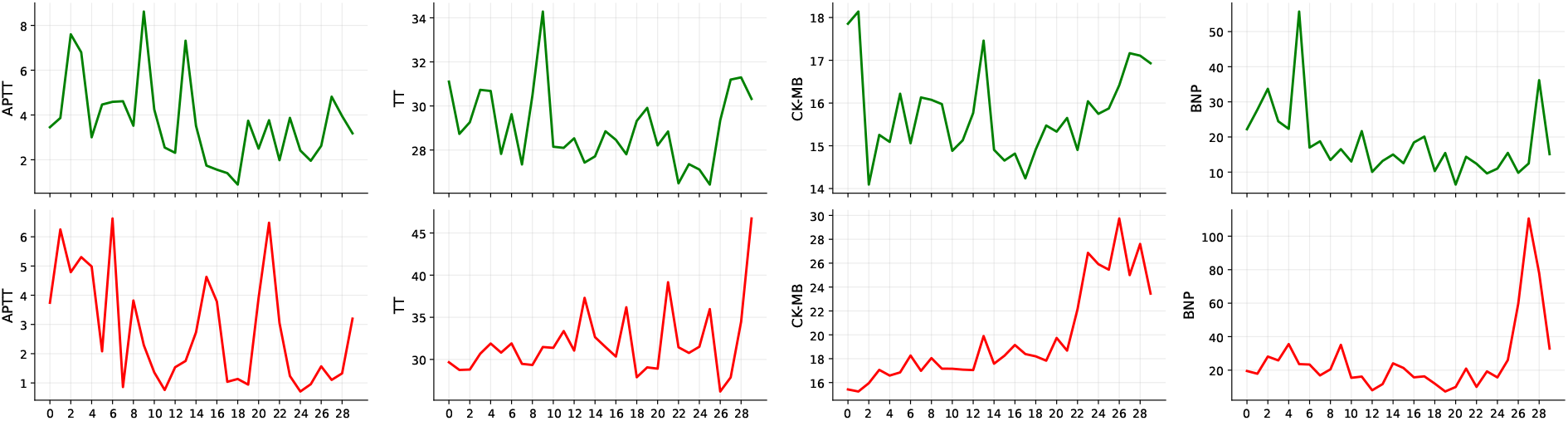
Visualization of synthetic cases generated with *λ* = 3 for four indicators: Positive Sample (Top Row) vs. Negative Sample (Bottom Row).

**Figure 3.8:**
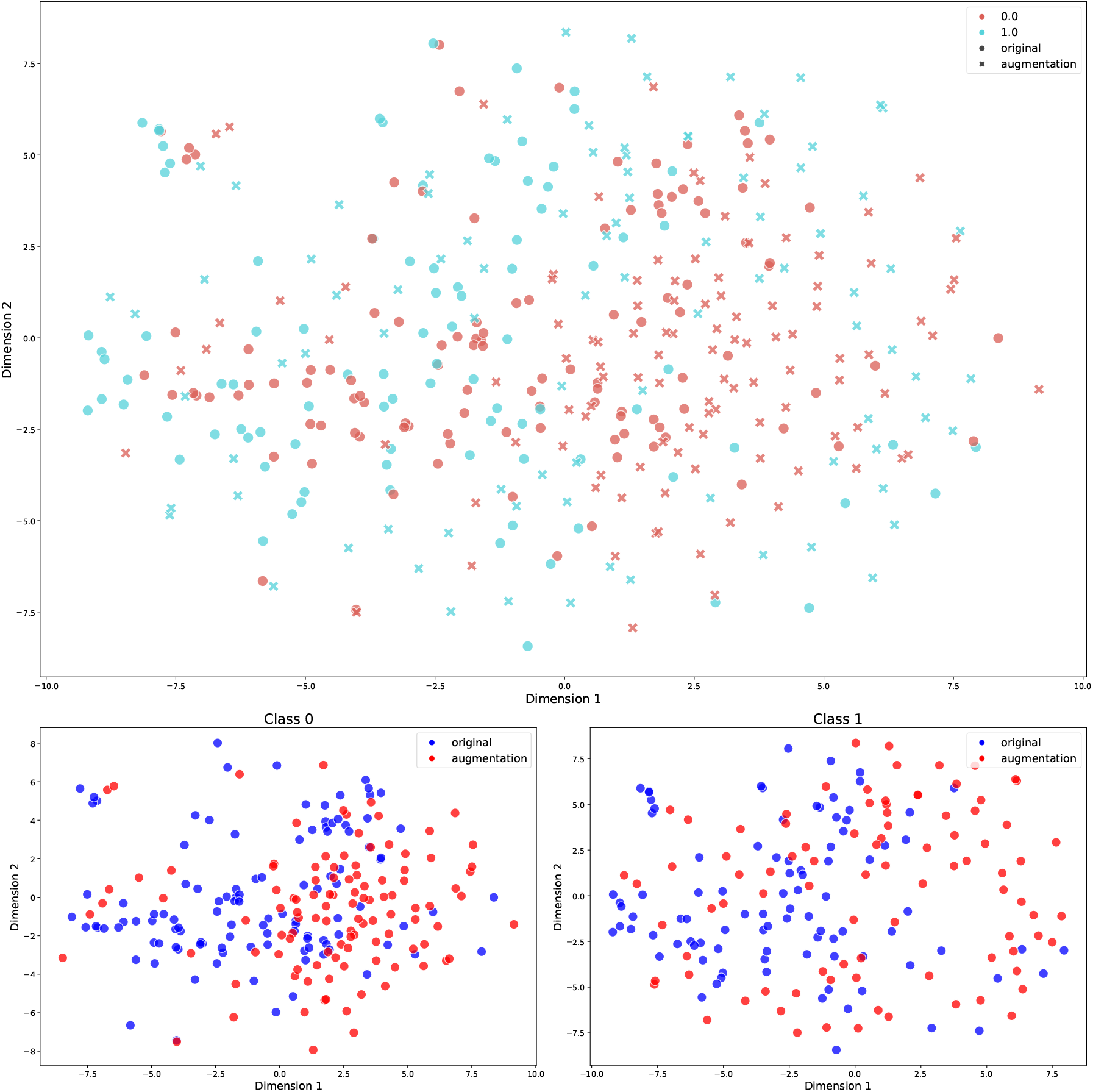
t-SNE Visualization of Original and Augmented Data for Class 0 and Class 1.

### 3.5 Reduced order modeling-Dimension reduction

#### 3.5.1 PCA

Principal Component Analysis (PCA) [1] is a widely used linear dimensionality reduction technique that identifies the directions in which a dataset exhibits the highest variance. By projecting the original variables onto these orthogonal axes, PCA converts correlated variables into a smaller set of uncorrelated components. This process not only reduces the dimensionality of high-dimensional data, but also reduces noise and redundant information, enabling more efficient downstream tasks such as classification.

Figure 3.9 displays the distribution of explained variance across the 31 principal components derived from the dataset. The green bars represent the explained variance ratio for each component, while the orange line indicates the cumulative explained variance. As shown, for the original dataset, the first 16 components account for 81.94% of the total variance, and the first 21 components account for 91.28%. For the augmented dataset, the first 18 components account for 81.95% of the variance, while the first 23 components account for 91.52%. We then select the components that account for 80% and 90% of the variance respectively, and train classification models using these reduced features. Detailed results are provided in Table 3.7.

**Figure 3.9:**
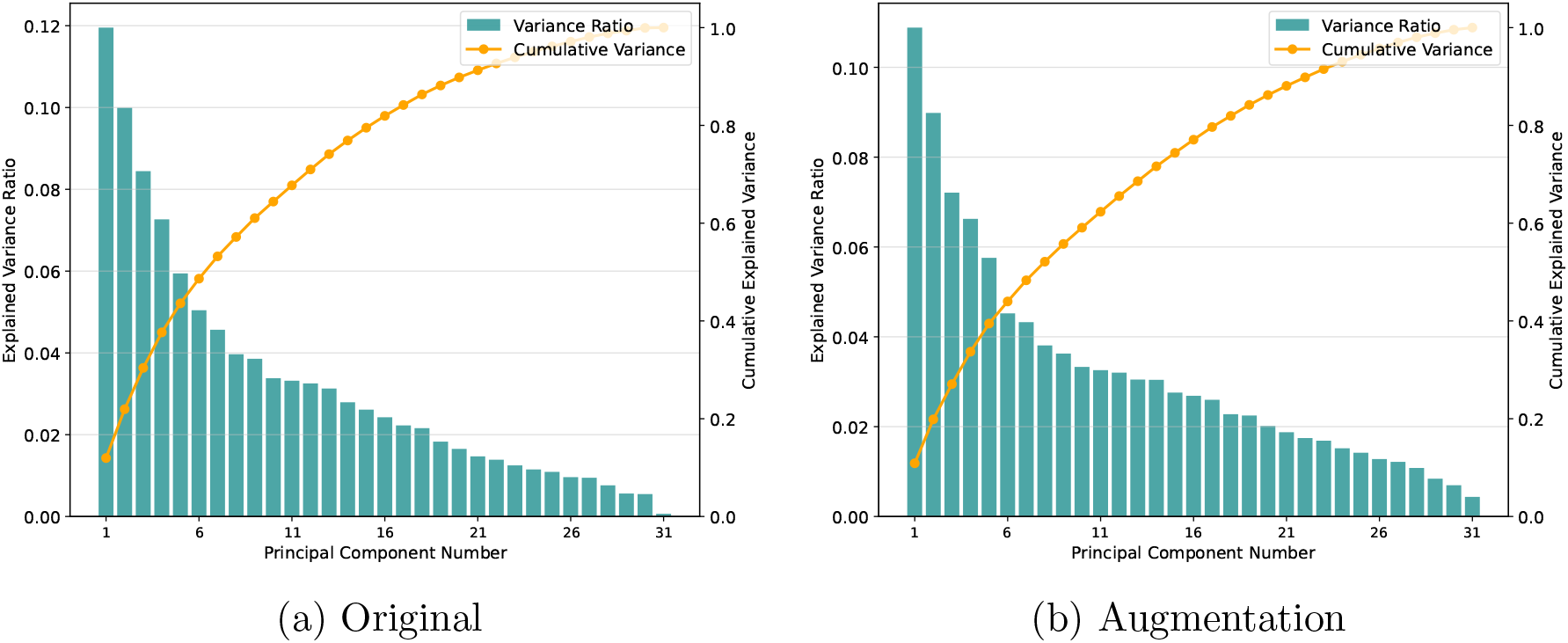
Explained variance distribution of principal components for the original and augmented Datasets. The green bars represent the explained variance ratio for each component, while the orange line indicates the cumulative explained variance.

**Table 3.7:** Classification performance under different PCA ratios for the original and augmented datasets.

| Dataset | PCA Ratio | Accuracy | Precision | Recall | F1 | AUROC |
| --- | --- | --- | --- | --- | --- | --- |
| Original | 80% | 82.86±2.19 | 82.18±2.78 | 89.58±3.54 | **85.65±1.83** | 87.45±2.06 |
| | 90% | **83.10±3.96** | **83.19±3.40** | 88.33±5.49 | 85.61±3.65 | **88.63±1.97** |
| | 100% | 82.62±3.56 | 81.64±3.73 | **90.00±3.51** | 85.56±2.86 | 87.36±1.16 |
| Augmentation | 80% | **86.19±2.19** | 83.85±2.41 | **94.17±5.96** | **88.56±2.28** | **89.14±2.71** |
| | 90% | 82.38±3.01 | 81.71±2.72 | 89.17±2.91 | 85.26±2.50 | 86.97±2.79 |
| | 100% | 84.76±3.76 | **84.23±4.06** | 90.42±2.81 | 87.18±3.04 | 86.02±1.44 |

Table 3.7 presents a comparison of classification metrics under two PCA variance thresholds (80% and 90%) alongside previous results for both the original and augmentation datasets. For the Original dataset, using the 90% ratio yields higher accuracy, precision, and AUROC compared to the previous results. In contrast, the augmentation dataset exhibits robust overall performance at the 80% ratio, with notably improved accuracy, recall, F1 and AUROC. These findings suggest that employing a PCA approach with either an 80% or 90% variance threshold can significantly enhance classification performance.

### 3.6 Feature Selection

#### 3.6.1 Original

We performed SHAP analysis to obtain the feature importance. According to Figure 3.10, we successively selected the top 1 to 7 indicators for each class, which are the same for both Class 0 and Class 1, and compared the resulting classification performance. The detailed results are presented in Table 3.8.

**Figure 3.10:**
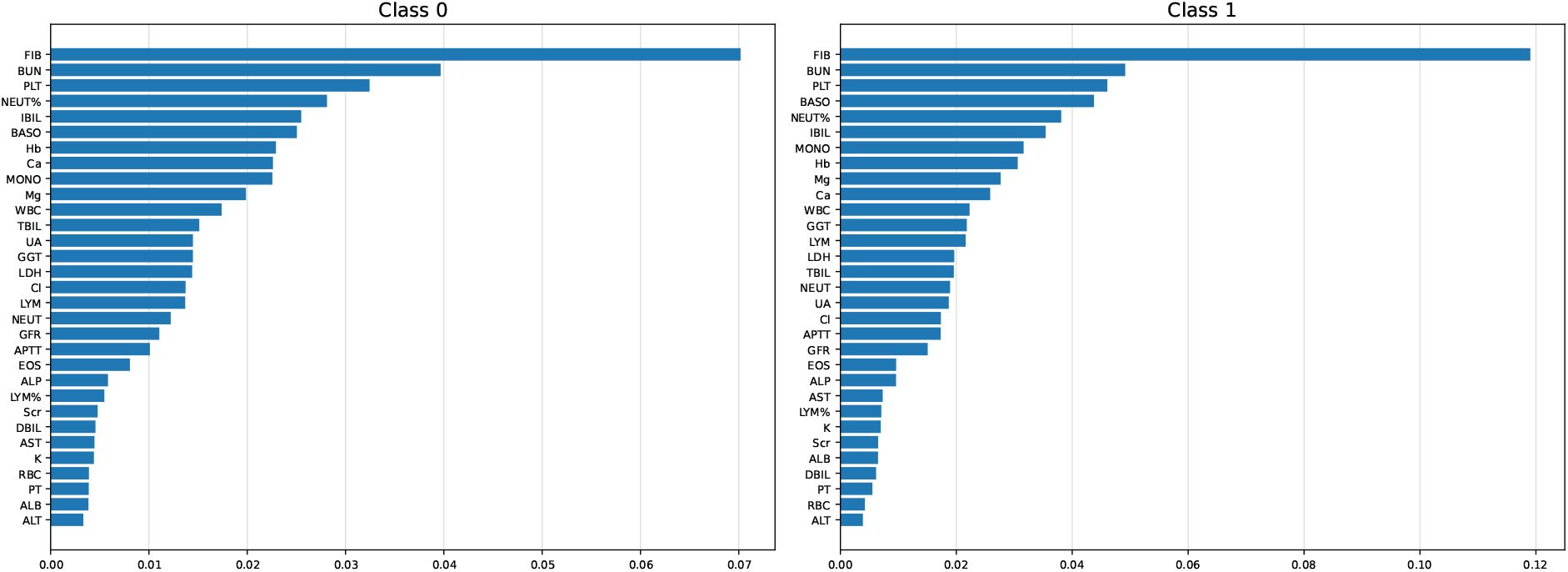
Feature importance rankings for Class 0 and Class 1.

**Table 3.8:** Comparison of performance using the top 1–7 indicators.

| Indicator Count | Accuracy | Precision | Recall | F1 | AUROC |
| --- | --- | --- | --- | --- | --- |
| 1 | 54.76±3.55 | 63.92±6.46 | 53.33±13.29 | 56.79±4.95 | 58.40±2.46 |
| 2 | 76.43±3.06 | 79.13±3.87 | 80.42±8.11 | 79.44±3.46 | 88.77±0.95 |
| 3 | 84.52±3.02 | 81.46±3.01 | 94.58±4.41 | 87.46±2.49 | 93.40±1.47 |
| 4 | 84.29±2.79 | 81.51±3.37 | 94.17±5.62 | 87.23±2.39 | 90.58±0.76 |
| 5 | 85.48±2.08 | 83.64±2.65 | 92.92±3.95 | 87.96±1.77 | 91.18±0.73 |
| 6 | 78.81±2.62 | 78.30±1.93 | 87.08±4.14 | 82.42±2.40 | 86.94±0.93 |
| 7 | 77.86±3.38 | 78.73±4.90 | 84.58±2.81 | 81.42±2.23 | 85.44±1.62 |
| 31 | 82.62±3.56 | 81.64±3.73 | 90.00±3.51 | 85.56±2.86 | 87.36±1.16 |

Table 3.8 presents the classification performance when successively selecting the top 1 to 7 indicators, as well as using all 31 features. With only one feature, the model achieves relatively low accuracy, but performance improves markedly with two features, which is comparable to the model trained on all 31 features. Notably, employing three features yields the highest accuracy, recall, F1 score and AUROC, outperforming the full model. These findings highlight the advantage of focusing on a smaller but more informative subset of indicators rather than using the entire feature set.

#### 3.6.2 Data Augmentation

Similarly, we selected the same top 1 to 7 indicators as the original dataset and compared the resulting classification performance. The detailed results are presented in Table 3.9.

**Table 3.9:**
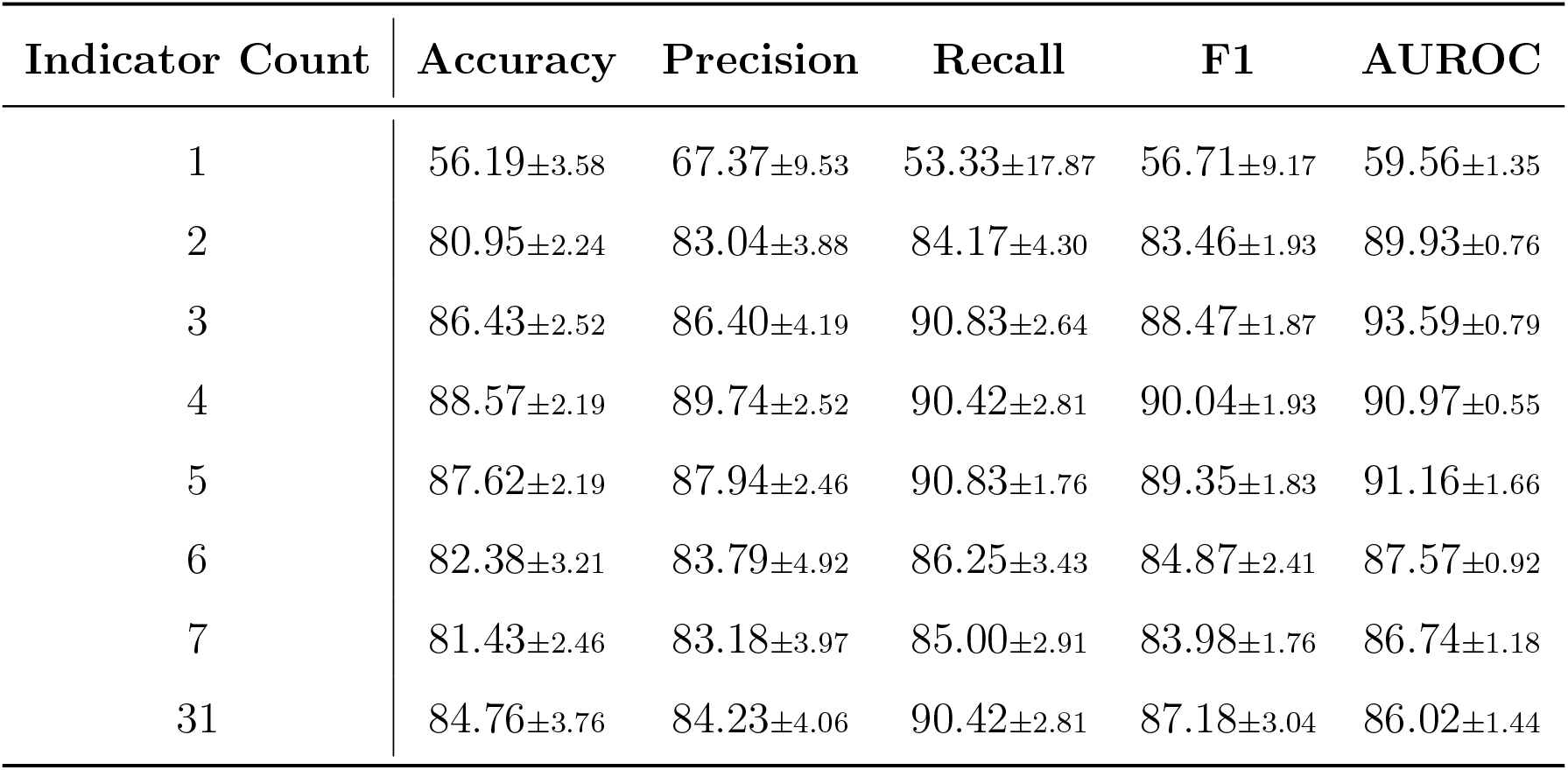
Comparison of performance using the top 1–7 indicators.

| Indicator Count | Accuracy | Precision | Recall | F1 | AUROC |
| --- | --- | --- | --- | --- | --- |
| 1 | 56.19±3.58 | 67.37±9.53 | 53.33±17.87 | 56.71±9.17 | 59.56±1.35 |
| 2 | 80.95±2.24 | 83.04±3.88 | 84.17±4.30 | 83.46±1.93 | 89.93±0.76 |
| 3 | 86.43±2.52 | 86.40±4.19 | 90.83±2.64 | 88.47±1.87 | 93.59±0.79 |
| 4 | 88.57±2.19 | 89.74±2.52 | 90.42±2.81 | 90.04±1.93 | 90.97±0.55 |
| 5 | 87.62±2.19 | 87.94±2.46 | 90.83±1.76 | 89.35±1.83 | 91.16±1.66 |
| 6 | 82.38±3.21 | 83.79±4.92 | 86.25±3.43 | 84.87±2.41 | 87.57±0.92 |
| 7 | 81.43±2.46 | 83.18±3.97 | 85.00±2.91 | 83.98±1.76 | 86.74±1.18 |
| 31 | 84.76±3.76 | 84.23±4.06 | 90.42±2.81 | 87.18±3.04 | 86.02±1.44 |

Table 3.9 presents the classification performance when successively selecting the top 1 to 7 indicators, compared to the baseline using all 31 features. The results observed in this dataset are consistent with those of the original dataset. Notably, the model utilizing the top four features outperforms the model trained on the full feature set. This finding highlights the advantage of focusing on a smaller but more informative subset rather than the complete feature set.

### 3.7 Interpretability-SHAP analysis

To better understand how the machine-learning model, TCN, makes decisions and to clarify importance of various features and time steps, we employ the SHAP analysis. SHapley Additive exPlanations (SHAP), introduced by Lundberg and Lee (2017) [18], is a game-theoretic approach designed for interpreting machine learning model predictions. It leverages Shapley values from cooperative game theory to quantify the contribution of each feature to model predictions, thereby providing transparent insights into the model’s decision-making processes. In this study, we selected the best-performing classification model based on CSDI’s results.

Figure 3.11 presents SHAP-based feature importance heatmaps for each class across 6 days. The color intensity indicates the mean absolute SHAP values, highlighting each feature’s contribution to model predictions over time. Notably, indicators such as FIB, BUN, and PLT show considerable importance at specific time steps, significantly influencing the classification results.

**Figure 3.11:**
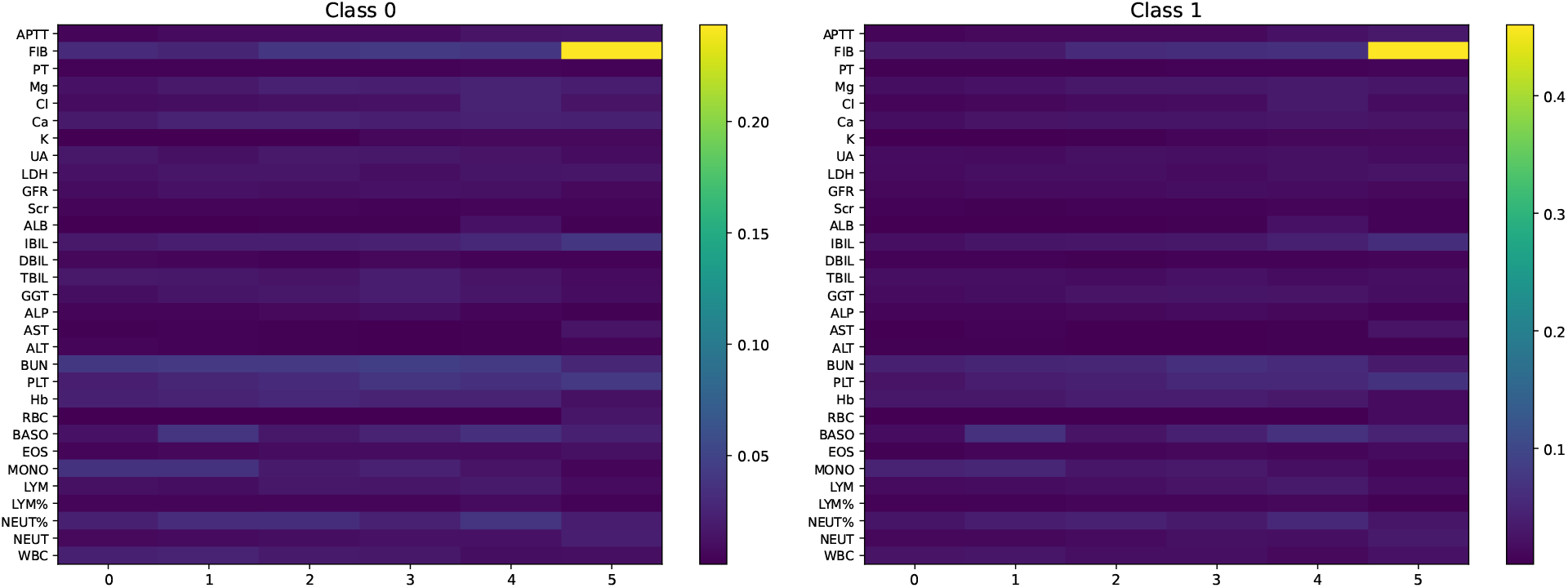
Heatmaps of feature importance across 6 days for Class 0 and Class 1 based on SHAP analysis.

Figure 3.12 illustrates the temporal trends in feature importance across 6 days for Class 0 and Class 1. Each time step shows mean absolute SHAP scores averaged across all features. For both classes, feature importance notably increases as the clinical outcome approaches, indicating that recent clinical measurements have a strong influence on classification results.

**Figure 3.12:**
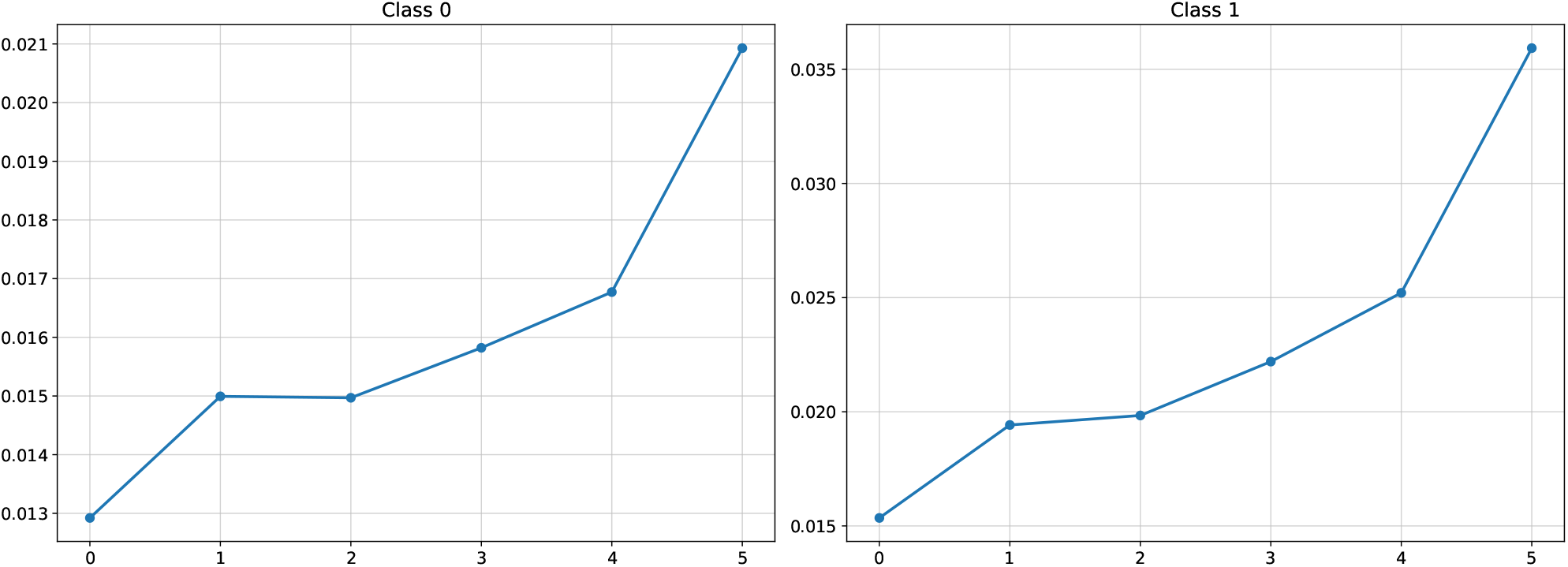
Trends in feature importance across 30 days for Class 0 and Class 1.

Figure 3.13 ranks feature importance based on mean absolute SHAP values averaged across all time steps. Features are presented in descending order, clearly highlighting those with the most significant contributions to model predictions. For Class 0, FIB, BUN, PLT and NEUT are the most critical features, while for Class 1, FIB, BUN, PLT and BASO top the list. These results provide valuable insights for clinicians about which indicators should pay more attention to.

**Figure 3.13:**
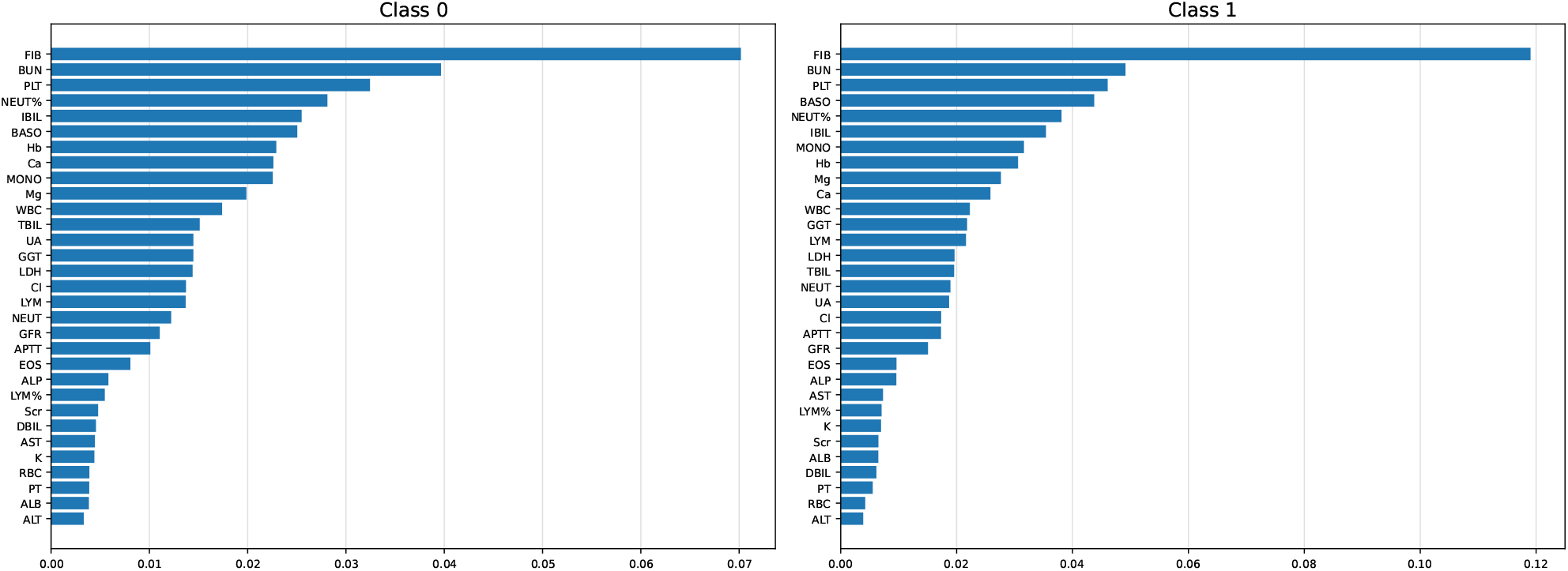
Feature importance rankings for Class 0 and Class 1.

Overall, these findings highlight the importance of temporal dynamics and feature selection in classification modeling. SHAP analysis not only enhances model interpretability but also identifies key features that can help clinical decision-making and potentially guide future model improvements.

### 3.8 Feature Selection + PCA

#### 3.8.1 Original

Based on the discussion above, we select the top 2-7 indicators and apply PCA to reduce their dimensionality to a single dimension. The results are presented in Table 3.10.

**Table 3.10:** Classification performance using the top 2–7 indicators, each reduced to a single dimension via PCA.

| Reduction (X→1) | Accuracy | Precision | Recall | F1 | AUROC |
| --- | --- | --- | --- | --- | --- |
| 2→1 | **75.48±1.61** | **77.54±1.51** | **80.42±2.81** | **78.92±1.52** | **85.16±1.48** |
| 3→1 | 63.81±7.51 | 69.52±7.79 | 71.25±20.74 | 68.17±9.30 | 68.43±9.21 |
| 4→1 | 63.57±4.05 | 70.50±10.51 | 71.67±19.42 | 68.38±5.51 | 68.82±9.03 |
| 5→1 | 63.10±4.38 | 70.32±9.92 | 69.58±19.55 | 67.40±6.38 | 66.30±7.76 |
| 6→1 | 57.86±5.62 | 64.37±7.90 | 69.17±26.22 | 63.01±12.57 | 56.16±7.09 |
| 7→1 | 61.19±4.05 | 68.43±7.37 | 67.50±22.72 | 65.06±8.55 | 64.05±4.90 |

Table 3.10 summarizes the classification performance when reducing the top 2–7 indicators to a single principal component. Notably, the model achieves its highest overall performance when two indicators are selected and reduced. Although this result falls slightly short of the performance achieved using the full feature set, it suggests that applying PCA to the selected features can further reduce dimensionality with only a marginal sacrifice in performance.

#### 3.8.2 Augmentation

We applied the same dimensionality reduction strategy to the augmented dataset, with results summarized in Table 3.11.

**Table 3.11:**
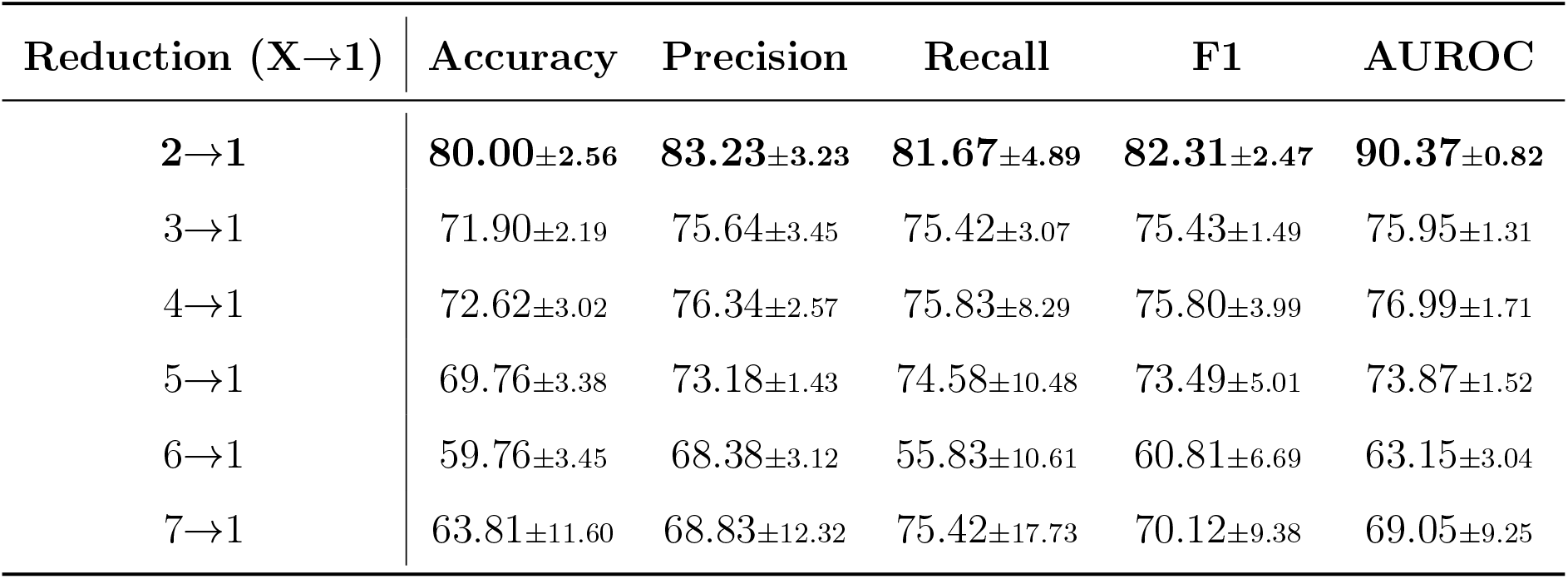
Classification performance using the top 2–7 indicators, each reduced to a single dimension via PCA.

Similar to the original dataset, the augmented model also achieves its highest performance using the top two indicators (2 → 1). Notably, this configuration is comparable to the model trained on the original two features, demonstrating that data augmentation enables PCA to significantly reduce dimensionality without sacrificing classification performance.

### 3.9 Ensemble Method

Based on our previous results, we selected three top-performing classifiers: the Random Forest classifier (*M*_1_), the PCA-based model preserving 80% of the variance (*M*_2_), and the model using the top-4 SHAP features (*M*_3_). To integrate their complementary strengths, we developed an ensemble method.

Specifically, we applied these three trained models to the training, validation, and test datasets to obtain their predicted probabilities. For any given input **x**, let *p*_*k*_ = *P*(*y* = 1 | **x**; *M*_*k*_) denote the probability output by the *k*-th classifier, where *k* = 1, 2, 3. We construct a three-dimensional vector **z** ∈ ℝ^3^ by concatenating these probabilities:

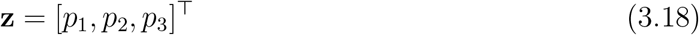

Subsequently, a Logistic Regression model [3] is trained on the training set. The final ensemble prediction *ŷ* is computed as:

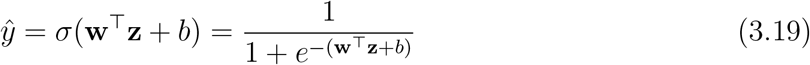

where **w** and *b* represent the learnable weights and bias, and *σ*(·) denotes the sigmoid function. As shown in Table 4.1, this ensemble method achieved superior performance, with all evaluation metrics surpassing 90%.

**Table 4.1:** Summary of classification performance across all methods. Models marked with ^*†*^ (Random Forest, PCA 80%, and SHAP top-4) are used in the ensemble.

| Method | Dataset | Accuracy | Precision | Recall | F1 | AUROC |
| --- | --- | --- | --- | --- | --- | --- |
| **Baseline classifiers** |  |  |  |  |  |  |
| TCN | Original | 82.62±3.56 | 81.64±3.73 | 90.00±3.51 | 85.56±2.86 | 87.36±1.16 |
| TCN | Augmented | 84.76±3.76 | 84.23±4.06 | 90.42±2.81 | 87.18±3.04 | 86.02±1.44 |
| **Traditional classifiers** |  |  |  |  |  |  |
| Random Forest<sup>†</sup> | Original | 87.14±2.43 | 93.61±3.20 | 83.33±3.73 | 88.09±2.34 | 88.52±0.95 |
| SVM | Original | 80.95±0.00 | 80.77±0.00 | 87.50±0.00 | 84.00±0.00 | 86.69±0.00 |
| XGBoost | Original | 85.24±2.08 | 93.25±2.20 | 80.00±3.12 | 86.08±2.10 | 87.87±0.84 |
| **Time-series-to-image** |  |  |  |  |  |  |
| Line | Original | 81.43±4.97 | 85.17±4.87 | 82.08±7.92 | 83.34±4.81 | 87.08±3.04 |
| Heatmap | Original | 80.00±3.72 | 81.05±2.58 | 85.00±7.50 | 82.78±3.96 | 86.30±2.07 |
| Bar | Original | 80.00±5.35 | 89.61±4.98 | 74.58±13.88 | 80.29±7.59 | 88.31±1.57 |
| **PCA on all indicators** |  |  |  |  |  |  |
| PCA 90% | Original | 83.10±3.96 | 83.19±3.40 | 88.33±5.49 | 85.61±3.65 | 88.63±1.97 |
| PCA 80%<sup>†</sup> | Augmented | 86.19±2.19 | 83.85±2.41 | 94.17±5.96 | 88.56±2.28 | 89.14±2.71 |
| **SHAP top-k indicators** |  |  |  |  |  |  |
| top-3 | Original | 84.52±3.02 | 81.46±3.01 | 94.58±4.41 | 87.46±2.49 | 93.40±1.47 |
| top-4<sup>†</sup> | Augmented | 88.57±2.19 | 89.74±2.52 | 90.42±2.81 | 90.04±1.93 | 90.97±0.55 |
| **SHAP top-k + PCA** |  |  |  |  |  |  |
| top-2 + PCA | Original | 75.48±1.61 | 77.54±1.51 | 80.42±2.81 | 78.92±1.52 | 85.16±1.48 |
| top-2 + PCA | Augmented | 80.00±2.56 | 83.23±3.23 | 81.67±4.89 | 82.31±2.47 | 90.37±0.82 |
| **Ensemble** | Original | 91.19±1.96 | 90.50±2.42 | 94.58±2.01 | 92.47±1.62 | 92.25±0.75 |

## 4 Results

We summarize all the classification results mentioned before in Table 4.1.

To evaluate the prognostic capabilities of the models, we modified the dataset by withholding the final day’s clinical records, thereby introducing a forecasting horizon. All previously described experiments were then repeated under this predictive setting. The corresponding results are presented in Table 4.2. As expected, excluding the most recent clinical measurements makes the task more challenging, resulting in a general decline in performance for most methods compared with the full six-day observation window. Nevertheless, our proposed ensemble approach exhibits remarkable robustness in this forecasting scenario, maintaining strong predictive performance with all evaluation metrics remaining above 85%.

**Table 4.2:** Summary of classification performance across all methods, where the final day’s clinical records are withheld. Models marked with ^*†*^ (Random Forest, PCA 80%, and SHAP top-5) are used in the ensemble.

| Method | Dataset | Accuracy | Precision | Recall | F1 | AUROC |
| --- | --- | --- | --- | --- | --- | --- |
| **Baseline classifiers** |  |  |  |  |  |  |
| TCN | Original | 79.05±3.13 | 81.17±2.42 | 82.50±4.73 | 81.77±3.01 | 85.90±1.83 |
| TCN | Augmented | 79.29±4.05 | 81.57±3.90 | 82.50±4.73 | 81.97±3.63 | 83.45±1.22 |
| **Traditional classifiers** |  |  |  |  |  |  |
| Random Forest<sup>†</sup> | Original | 86.43±1.52 | 93.43±2.08 | 82.08±2.67 | 87.35±1.51 | 88.38±1.42 |
| SVM | Original | 83.33±0.00 | 86.96±0.00 | 83.33±0.00 | 85.11±0.00 | 85.47±0.08 |
| XGBoost | Original | 83.57±2.70 | 92.31±3.81 | 77.92±3.75 | 84.41±2.52 | 87.48±1.39 |
| **Time-series-to-image** |  |  |  |  |  |  |
| Line | Original | 77.86±4.13 | 81.48±3.79 | 79.58±7.56 | 80.28±4.33 | 86.23±2.72 |
| Heatmap | Original | 69.76±6.74 | 70.70±4.70 | 80.42±11.49 | 74.93±6.78 | 78.82±6.51 |
| Bar | Original | 83.81±2.56 | 81.29±2.76 | 93.33±3.82 | 86.82±2.07 | 88.26±1.55 |
| **PCA on all indicators** |  |  |  |  |  |  |
| PCA 80%<sup>†</sup> | Original | 82.86±2.70 | 85.47±3.70 | 84.58±3.95 | 84.93±2.36 | 87.78±1.99 |
| PCA 80% | Augmented | 81.90±3.01 | 80.77±3.63 | 90.00±4.03 | 85.04±2.41 | 86.90±1.61 |
| **SHAP top-k indicators** |  |  |  |  |  |  |
| top-6 | Original | 80.48±3.33 | 83.09±3.80 | 82.92±4.14 | 82.91±2.82 | 89.58±1.41 |
| top-5<sup>†</sup> | Augmented | 82.86±3.13 | 86.05±3.82 | 83.75±4.14 | 84.80±2.85 | 89.42±1.47 |
| **SHAP top-k + PCA** |  |  |  |  |  |  |
| top-7 + PCA | Original | 66.19±6.02 | 70.24±8.08 | 76.67±17.15 | 71.53±7.01 | 74.63±6.77 |
| top-7 + PCA | Augmented | 67.86±4.09 | 73.20±3.60 | 70.00±12.85 | 70.79±6.58 | 76.18±1.64 |
| **Ensemble** | Original | 89.52±1.66 | 93.43±2.09 | 87.92±3.07 | 90.54±1.59 | 88.94±1.14 |

## 5 Discussions

This study presents a comprehensive machine-learning framework for prognostic modeling in ICU patients using routine biochemical data. Our investigation systematically addressed the challenges of missing data, model selection, and the constraints of a small clinical dataset. To overcome these issues, we employed a multi-faceted strategy that incorporated advanced data imputation, rigorous classifier benchmarking, data augmentation, and dimensionality reduction. The models achieved clinically acceptable performance, with accuracy and precision metrics generally exceeding 85% and AuROC surpassing 90% in certain configurations.

A foundational component of our framework was addressing missing values, which is a pervasive issue in clinical datasets. Our results confirm that machine learning-based imputation methods significantly outperform traditional approaches like mean or median imputation. Notably, the conditional score-based diffusion model (CSDI) yielded the lowest imputation error (MSE=0.252, MAE=0.190). The superiority of this imputation strategy was further validated in downstream tasks, where data imputed by CSDI led to the highest classification accuracy when paired with the TCN model. This highlights the critical importance of a robust imputation strategy, as it directly impacts the quality of data fed into classification models and, consequently, their performance.

In the classification task, a benchmark of various models revealed that while the TCN deep learning model performed well, it was surpassed by other approaches. Traditional machine learning models, particularly Random Forest, demonstrated superior overall performance. Furthermore, the exploration of a novel time-series-to-image encoding method proved to be a highly effective strategy. The model using a bar plot representation not only outperformed the TCN baseline, but also achieved the highest precision (89.61 *±* 4.98) among all tested classifiers. This suggests that for time-series data from small cohorts, traditional models and novel representation techniques that leverage powerful pre-trained vision models can be more effective than deep learning sequence models.

To address the limitations of a small patient cohort, we employed two distinct strategies: generative data augmentation and dimensionality reduction. First, we utilized a conditional diffusion model to generate additional 200 synthetic patient records. This data augmentation improved performance across all evaluation metrics, notably increasing accuracy by 2.59%. Conversely, reducing the data’s dimensionality also enhanced model performance. For instance, applying Principal Component Analysis (PCA) to retain 16-21 of the data’s principal components improved classification results, while selecting just the top seven indicators via SHAP analysis yielded higher performance than using the entire feature set. These findings demonstrate that for small, high-dimensional clinical datasets, performance can be significantly enhanced by either synthetically expanding the data or by using dimensionality reduction strategies to identify the most informative features.

However, this study has several limitations. First, as a retrospective analysis based on a small cohort of 332 patients, these findings require external validation on a larger dataset to ensure generalizability. Second, the interpretability analysis was performed on the TCN model, which was not the best-performing classifier. Therefore, future work should apply SHAP analysis to the superior ViTST models to better identify clinically relevant features. Finally, future work should also focus on exploring additional time-series-to-image encoding methods and further refining both the imaging and generative modeling techniques. Rigorous external validation across diverse cohorts will be essential to confirm the framework’s effectiveness and enhance its utility for clinical decision support.

## 6 Conclusion

This study presents a comprehensive framework for developing and validating machine-learning prognostic models for ICU patients, leveraging routine biochemical indicators from a small patient cohort. This comprehensive framework successfully integrated advanced data imputation using diffusion models, data augmentation to expand the dataset, and dimensionality reduction via both Principal Component Analysis (PCA) and SHAP-based feature selection. Overall, the developed models achieved clinically acceptable performance, with accuracy and precision rates above 85% and an AUROC exceeding 90% in some optimized configurations, demonstrating a robust toolkit for prognostication.

This work demonstrates that the CSDI diffusion model provides superior data imputation, which in turn improves downstream classification tasks. A comparative benchmark of classifiers reveals that traditional models like Random Forest and novel time-series-to-image encoding approaches can outperform a baseline deep learning sequence model for this classification task. Furthermore, the results establish that both generating synthetic patient data and reducing the feature set to a few key indicators lead to significant improvements in predictive performance.

However, the study has several limitations. The generalizability of the findings is limited by the small cohort size, which necessitates future validation on larger, more diverse datasets. Furthermore, the interpretability analysis was performed on the TCN model, which was not the best-performing classifier, potentially affecting the conclusions about feature importance. Finally, while the novel time-series-to-image approach proved effective, further investigation into a broader range of encoding strategies is warranted.

This work successfully demonstrates the feasibility of building robust prognostic tools from small and incomplete clinical datasets. It provides a comprehensive and modular foundation that can be readily deployed to aid physicians and integrated into future digital twin technologies for specific diseases.

## Data Availability

All data produced in the present study are available upon reasonable request to the authors

https://sc.edu/study/colleges_schools/artsandsciences/mathematics/our_people/directory/wang_qi.php

## Appendix

### 6.1 Confusion matrix and evaluation metrics

The confusion matrix is defined in Table 6.1. This matrix summarizes the counts of correct and incorrect predictions by comparing the ground truth labels with the model’s predictions. It consists of the following elements:

**Table 6.1:** Confusion Matrix.

|  | **Predicted Positive** | **Predicted Negative** |
| --- | --- | --- |
| **Actual Positive** | TP | FN |
| **Actual Negative** | FP | TN |

- **True Positive (TP)**: the number of positive instances correctly predicted as positive.
- **False Negative (FN)**: the number of positive instances incorrectly predicted as negative.
- **False Positive (FP)**: the number of negative instances incorrectly predicted as positive.
- **True Negative (TN)**: the number of negative instances correctly predicted as negative.

From this confusion matrix, several commonly used metrics are derived:

- Accuracy measures the proportion of instances that are correctly classified:

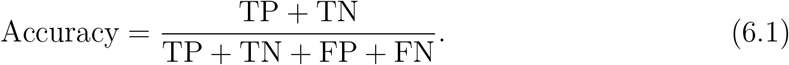
- Precision measures the proportion of predicted positive instances that are truly positive:

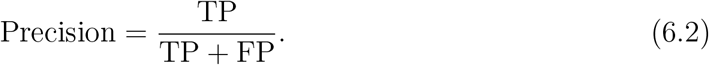
- Recall measures the proportion of actual positive instances that are correctly identified:

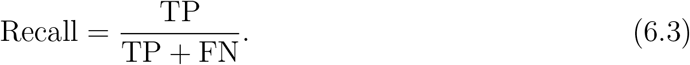
- The F1-Score is the harmonic mean of Precision and Recall, providing a balanced measure of both:

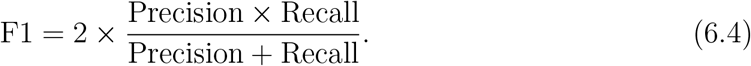
- The ROC (Receiver Operating Characteristic) curve plots the True Positive Rate (TPR) against the False Positive Rate (FPR) across various threshold settings. The AUROC is defined as the area under this curve.

## Acknowledgments

The authors would like to acknowledge the contribution made by Dr. Hua Jiang’s group to this study by sharing with us their dataset collected. Xiang Cao, Jianguo Hou, Xinhang Wei, and Qi Wang’s research is partially supported by NSF award OIA-2242812; Qi Wang’s research is partially supported by an SC GAIN-CRP award. The funders played no role in study design, data collection, analysis and interpretation of data, or the writing of this manuscript.

## Contributions

X.C. and Q. W. conceived the study. X.C. conceptualized the model and conducted the analyses. J.H. and X. W. performed the literature review to inform model parameters. X.C., J.H., and X.W. validated the model structure and assumptions and provided feedback on parameter selection. X.C. and Q.W. interpreted the results and prepared the first draft. Q.W. and C.C. revised the manuscript. Q.W. secured funding for the project. All authors contributed to the interpretation of the findings, reviewed the manuscript, and approved the final manuscript.

## Data availability

The datasets used and/or analyzed during the current study are available from the corresponding author on reasonable request.

## Code availability

The underlying code for this study and training/validation datasets is not publicly available but may be made available to researchers on reasonable requests from the corresponding authors.

## References

[1] Hervé Abdi and Lynne J Williams. Principal component analysis. Wiley interdisciplinary reviews: computational statistics, 2(4):433–459, 2010.

[2] Shaojie Bai, J Zico Kolter, and Vladlen Koltun. An empirical evaluation of generic convolutional and recurrent networks for sequence modeling. arXiv preprint arXiv:1803.01271, 2018.

[3] Christopher M Bishop and Nasser M Nasrabadi. Pattern recognition and machine learning, volume 4. Springer, 2006.

[4] Leo Breiman. Random forests. Machine learning, 45:5–32, 2001.

[5] Tianqi Chen and Carlos Guestrin. Xgboost: A scalable tree boosting system. In Proceedings of the 22nd acm sigkdd international conference on knowledge discovery and data mining, pages 785–794, 2016.

[6] M. Z. I. Chowdhury and T. C. Turin. Variable selection strategies and its importance in clinical prediction modelling. Fam Med Community Health, 8(1):e000262, 2020.

[7] G. S. Collins, J. B. Reitsma, D. G. Altman, and K. G. Moons. Transparent reporting of a multivariable prediction model for individual prognosis or diagnosis (tripod): the tripod statement. BMJ, 350:g7594, 2015.

[8] Hong-Fei Deng, Ming-Wei Sun, Yu Wang, Jun Zeng, Ting Yuan, Ting Li, Di-Huan Li, Wei Chen, Ping Zhou, Qi Wang, and Hua Jiang. Evaluating machine learning models for sepsis prediction: A systematic review of methodologies. iScience, 25:103651, 2022.

[9] Prafulla Dhariwal and Alexander Nichol. Diffusion models beat gans on image synthesis. Advances in neural information processing systems, 34:8780–8794, 2021.

[10] W. A. Farooqui, M. Uddin, R. Qadeer, and K.d Shafique. Latent class trajectories of biochemical parameters and their relationship with risk of mortality in icu among acute organophosphorus poisoning patients. Sci Rep, 12(1):11633, 2022.

[11] Vincent Fortuin, Dmitry Baranchuk, Gunnar Rätsch, and Stephan Mandt. Gp-vae: Deep probabilistic time series imputation. In International conference on artificial intelligence and statistics, pages 1651–1661. PMLR, 2020.

[12] E. Ghaznavi-Rad, M. Khosravi, and M. Sayyadi. The importance of using routine laboratory tests in the diagnosis and prognosis of patients with coronavirus disease 2019: Shedding light on clinical laboratory data in covid-19. J Clin Lab Anal, 36(11):e24713, 2022.

[13] Marti A. Hearst, Susan T Dumais, Edgar Osuna, John Platt, and Bernhard Scholkopf. Support vector machines. IEEE Intelligent Systems and their applications, 13(4):18–28, 1998.

[14] Jonathan Ho and Tim Salimans. Classifier-free diffusion guidance. arXiv preprint arXiv:2207.12598, 2022.

[15] Seto W. K., C. F. Lee, C. L. Lai, P. P. Ip, D. Y. Fong, J. Fung, D. K. Wong, and M. F. Yuen. A new model using routinely available clinical parameters to predict significant liver fibrosis in chronic hepatitis b. PLoS One, 6(8):e23077, 2011.

[16] M Kristensen, AKS Iversen, TA Gerds, R Østervig, JD Linnet, C Barfod, KHW Lange, G Sölétormos, JL Forberg, J Eugen-Olsen, LS Rasmussen, M Schou, L Køber, and K. Iversen. Routine blood tests are associated with short term mortality and can improve emergency department triage: a cohort study of ¿12,000 patients. Scand J Trauma Resusc Emerg Med., 25(1):115, 2017.

[17] Zekun Li, Shiyang Li, and Xifeng Yan. Time series as images: Vision transformer for irregularly sampled time series. Advances in Neural Information Processing Systems, 36:49187–49204, 2023.

[18] Scott M Lundberg and Su-In Lee. A unified approach to interpreting model predictions. Advances in neural information processing systems, 30, 2017.

[19] Xiaoye Miao, Yangyang Wu, Jun Wang, Yunjun Gao, Xudong Mao, and Jianwei Yin. Generative semi-supervised learning for multivariate time series imputation. In Proceedings of the AAAI conference on artificial intelligence, volume 35, pages 8983–8991, 2021.

[20] Yang Song, Jascha Sohl-Dickstein, Diederik P Kingma, Abhishek Kumar, Stefano Ermon, and Ben Poole. Score-based generative modeling through stochastic differential equations. arXiv preprint arXiv:2011.13456, 2020.

[21] Yusuke Tashiro, Jiaming Song, Yang Song, and Stefano Ermon. Csdi: Conditional score-based diffusion models for probabilistic time series imputation. Advances in Neural Information Processing Systems, 34:24804–24816, 2021.

[22] Laurens Van der Maaten and Geoffrey Hinton. Visualizing data using t-sne. Journal of machine learning research, 9(11), 2008.

[23] A Vaswani. Attention is all you need. Advances in Neural Information Processing Systems, 2017.

